# Longitudinal associations of sustained low or high income and income variability with incident cardiovascular disease in individuals with type 2 diabetes: a retrospective population-based cohort study

**DOI:** 10.1101/2023.09.29.23296223

**Authors:** Yong-Moon Mark Park, Jong Ha Baek, Hong Seok Lee, Tali Elfassy, Clare C. Brown, Mario Schootman, Marie-Rachelle Narcisse, Seung-Hyun Ko, Pearl A. McElfish, Michael R. Thomsen, Benjamin C. Amick, Seong-Su Lee, Kyungdo Han

**Affiliations:** Department of Epidemiology, Fay W. Boozman College of Public Health, University of Arkansas for Medical Sciences, Little Rock, AR; Winthrop P. Rockefeller Cancer Institute, University of Arkansas for Medical Sciences, Little Rock, AR; Department of Internal Medicine, Gyeongsang National University Changwon Hospital, Gyeongsang National University College of Medicine, Changwon, South Korea; University of Arizona, Department of Medicine, Tucson, AZ; Department of Public Health Sciences, University of Miami Miller School of Medicine, Miami, FL; Department of Health Policy and Management, Fay W. Boozman College of Public Health, University of Arkansas for Medical Sciences, Little Rock, AR; Department of Medicine, College of Medicine, University of Arkansas for Medical Sciences, Little Rock, AR; Division of Endocrinology and Metabolism, Department of Internal Medicine, St. Vincent’s Hospital, College of Medicine, The Catholic University of Korea, Seoul, South Korea; Division of Endocrinology and Metabolism, Department of Internal Medicine, College of Medicine, The Catholic University of Korea, Seoul, South Korea; Department of Statistics and Actuarial Science, Soongsil University, Seoul, South Korea

**Keywords:** type 2 diabetes, income dynamics, cardiovascular disease, risk factor

## Abstract

**Background and Aims:** Longitudinal change in income is crucial in explaining cardiovascular health inequalities, but there is limited evidence for cardiovascular disease (CVD) risk associated with income dynamics over time among individuals with type 2 diabetes (T2D).

**Methods:** Using a nationally representative sample from the Korean Health Insurance Service database, 1,528,108 adults with T2D aged 30-64 years and no history of CVD were enrolled between 2009-2012 (mean follow-up of 7.3 years). Using monthly health insurance premiums information, income levels were assessed annually for 4 years before the baseline year. Income variability was defined as the intraindividual standard deviation of the percent change in income across 5 years. The primary outcome was a composite event of incident fatal and nonfatal CVD (myocardial infarction, stroke, and heart failure) using insurance claims. Hazard ratios (HRs) and 95% confidence intervals (CIs) were estimated after adjusting for potential confounders.

**Results:** Sustained low income (i.e., lowest income quartile) over 5 years was associated with increased CVD risk (HR_n=5years vs. n=0years_ 1.38, 95% CI 1.35-1.41; P_trend_<0.0001), whereas sustained high income (i.e., highest income quartile) was associated with decreased CVD risk (HR 0.71_n=5years vs. n=0years_ 95% CI 0.70-0.72; P_trend_<0.0001). High-income variability was associated with increased CVD risk (HR_highest vs. lowest quartile_ 1.25, 95% CI 1.22-1.27; P_trend_<0.001). Individuals who experienced an income decline across 5 years leading up to baseline had increased CVD risk, particularly in a decrease to the lowest income level (i.e., Medical Aids beneficiaries), regardless of initial income status. Sensitivity analyses, including potential mediators, such as lifestyle-related factors and obesity, supported the results.

**Conclusions:** Among non-elderly Korean adults with T2D, sustained low income, higher income variability, and income declines were associated with increased CVD risk. Our findings highlight the need to understand better the mechanisms by which income dynamics impact CVD risk among individuals with T2D.

## INTRODUCTION

Individuals with type 2 diabetes (T2D) are at increased risk of cardiovascular disease (CVD).^1^ Low socioeconomic status (SES), based on income, educational attainment, and employment status,^2^ confers elevated CVD risk as much as traditional cardiovascular (CV) risk factors such as age, hypertension, dyslipidemia, and lifestyle factors.^3^ In those with T2D, low SES is associated with non-attainment of optimal diabetes care (glycemic, lipid, and blood pressure control, adherence to healthy behaviors, and weight control),^4–6^ leading to an elevated risk of micro- and macro-vascular complications,^7–9^ and CV-related and all-cause mortality.^10,11^ The adverse health effects of low SES may be more substantial in non-elderly individuals with T2D who are in the economically active age group and more affected by comorbidities and CV risk factors associated with increased mortality risk than in older adults.^12^

Although low income is associated with increased CVD risk in those with T2D,^7^ it is unclear whether low income plays a causal role or is just a surrogate indicator for other behavioral and clinical characteristics contributing to elevated CVD risk.^13^ Evidence on income status and CVD risk among individuals with T2D has been limited by cross-sectional study designs in which reverse causality cannot be ruled out;^4,14–18^ use of group- or area-level income data,^8,15–18^ which may be prone to the ecological fallacy;^19^ and lack of the evaluation of potential mediators, such as smoking, alcohol consumption, physical inactivity, and obesity.^20^ Furthermore, prior studies used single-point individual income measures,^9,11,18,21,22^, which ignores income stability and variability over time.^23^ Longitudinal change in income may be crucial in explaining CV health inequalities.^24^ Similarly, single-point exposure measures may underestimate CVD risk as income volatility and drops have increased CVD risk in the general population.^13,25,26^

In the present study, we used data from the Korean National Health Insurance System (NHIS) to examine the longitudinal association of sustained low- or high-income status, income variability, and changes in income with incident fatal and nonfatal CVD in non-elderly individuals with T2D.

## METHODS

### Data Sources

The NHIS, a single-payer public health insurance system for all residents in South Korea, covers 97% of the population (∼50 million individuals). The Medical Aid program covers the remaining 3% of the population and provides medical benefits to those with the lowest income (less than $600 monthly).^27^ The NHIS database includes all claims-based information, including patient demographics, drug prescriptions, International Classification of Disease, Tenth Revision (ICD-10) diagnostic codes, insurers’ payment coverage, patients’ deductions, and claimed treatment details.^28,29^ The NHIS also provides standardized national health screening examinations biannually or before employment, which collects data on health behaviors, anthropometric measurements, and laboratory tests. This study was approved by the Institutional Review Board of Soongsil University, Seoul, South Korea, and complied with the ethical guidelines of the World Medical Association Declaration of Helsinki.

### Study Population

We included 1,909,492 individuals with T2D who underwent the national health screening examinations from 2009 through 2012 (baseline year). The study was limited to individuals between the ages of 30 and 64 years at baseline. We decided on 30 years as a lower age limit because most Korean men are precluded from economically active states while in military service in their twenties, in addition to reducing the potential misclassification of those with type 1 diabetes as T2D.^30^ The presence of T2D was defined as either having ≥1 claim per year for an anti-diabetic medication prescription, ICD-10 codes corresponding to T2D (E11-14), or a high fasting glucose level (≥126 mg/dL). Individuals with type 1 diabetes (E10) or gestational diabetes mellitus (O24) were excluded.

### Definition of income status

Because the NHIS does not record household income data, we used monthly health insurance premiums, which are unchanging unless there is a significant change in income, as a proxy for household income.^31^ Wage income among employee insured (government employees, school employees, and industrial workers) and self-employed income with the household property value (for self-employed) determines health insurance premiums.^32^ Using the health insurance premium given as 20 quantile levels, we divided annual income status into 4 levels (quartiles), following a previous study.^33^ Individuals in the lowest income quartile and Medical Aid program beneficiaries were categorized as a low-income group, and those in the highest income quartile were classified as a high-income group. Individual income data were collected from the baseline year backward 4 years to establish a pre-baseline income status (Fig 1). We used the following definitions of income parameters.

1. <u>Cumulative number of years being in the low- or high-income group</u> was counted during the 4 years before the baseline year. For example, if an individual enrolled in 2010 was in the low-income group consecutively from 2007 to 2009 but not in 2006 and 2010, the cumulative years of low-income status were recorded as 3.
2. <u>Income variability</u> was calculated using 20 income quantiles analyzed as a continuous variable by assigning consecutive integers to each quantile to summarize income fluctuations over the study period. Income variability was defined as the intraindividual standard deviation (SD) of the percent change in income. We first calculated the percent change between every 2 consecutive years as [(Y_t2_–Y_t1_)/0.5 (Y_t1_+Y_t2_)]×100 (Y=income, t=time),^25^ and then calculated the SD of those percent changes by the individual. The SD for each individual was categorized into quartiles to define groups based on income variability.
3. Individuals’ <u>baseline income status</u> was categorized into quartiles. Medical Aid beneficiaries, a part of Quartile 1, were classified separately.
4. Additionally, <u>changes in income status (i.e., rise and decline)</u> were compared between the first assessment (4 years before baseline) and the last (baseline).

**Figure 1.**
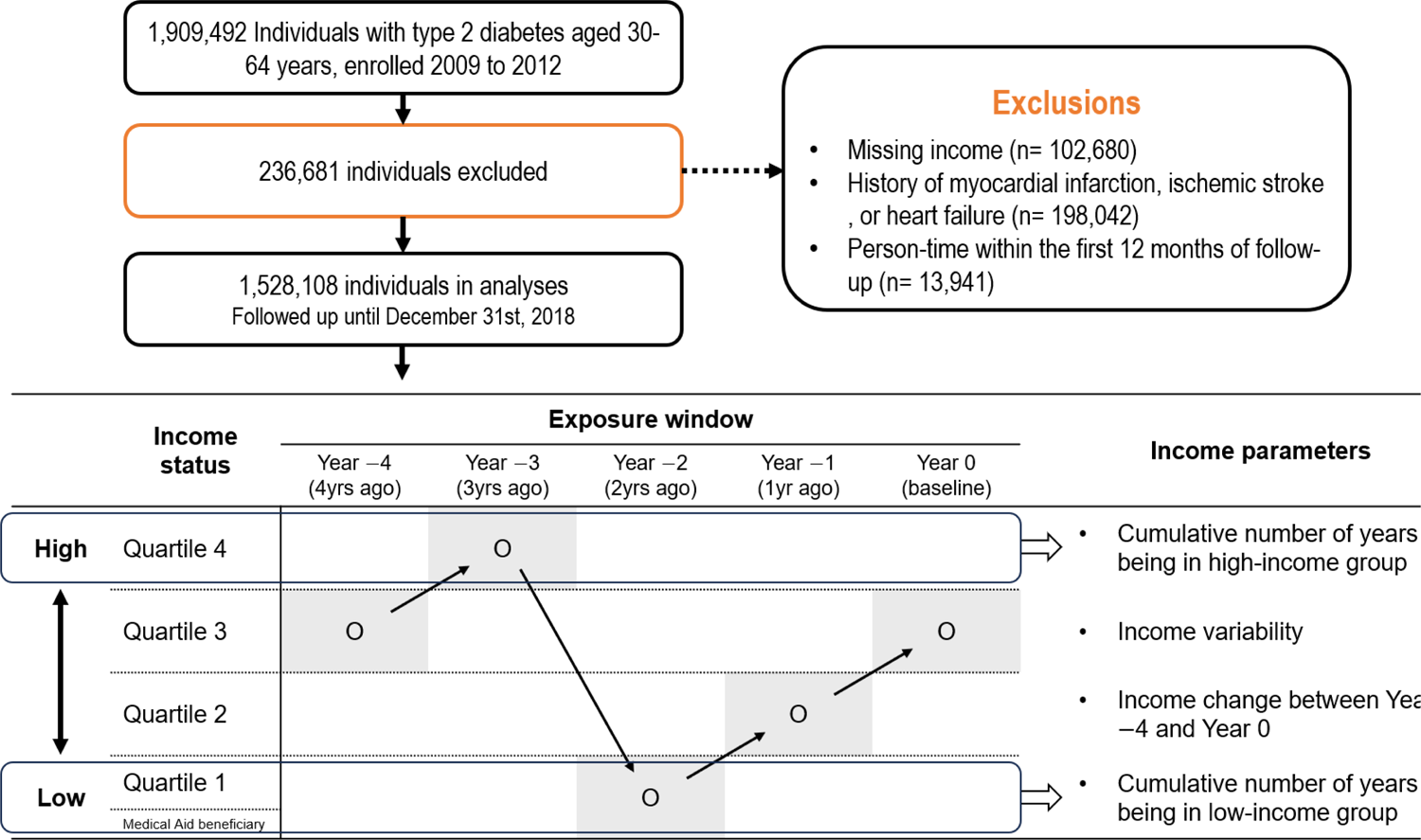
Study population flow diagram and income parameters Cumulative number of years being in the low- or high-income group was counted during the 4 years before the baseline year. Income variability was calculated using 20 income quantiles analyzed as a continuous variable to summarize income fluctuations over the same study period. Changes in income status (i.e., rise and decline) were compared between the first assessment (4 years before baseline) and the last (baseline), where Medical Aid beneficiaries, a part of Quartile 1, were classified separately.

### Ascertainment of CVD and covariates

Individuals’ primary outcome was the composite of multiple CVD outcomes, which were defined as the first occurrence of one of the following ICD-10 codes: MI (I21-22), stroke (I63.54), HF (I50), or CV mortality.^28^ Incident CVD outcomes were identified for each individual up to December 31, 2018. Covariates included sociodemographic factors, CV risk factors, health behaviors, and T2D duration and treatments (see **Supplemental methods** for details).^2,34^

### Statistical analysis

We excluded those with missing income data (n=102,680); with a previous history of myocardial infarction (MI), ischemic stroke, or heart failure (HF) (n=198,042); or with at least 1 missing covariate (n=66,721). Additionally, individuals with a CVD diagnosis during their first year of follow-up were excluded to reduce potential bias related to undetected CVD present at baseline (n=13,941). In all, 1,528,108 individuals were included in the analysis (**Fig 1**). Person-time was calculated from the individual’s age 1 year after baseline until the age of CVD diagnosis or until death, last follow-up, or whichever occurred first.

Baseline characteristics were presented as counts and percentages or as means with SDs. The CVD incidence rate was computed by dividing the number of events by the total number of person-years of follow-up presented per 1,000 person-years. Cox proportional hazards models were applied to estimate hazard ratios (HRs) and 95% confidence intervals (CIs) for the association between income dynamics and CVD risk. The proportional hazards assumption was checked using the Schoenfeld residuals test with the logarithm of the cumulative hazards function for income parameters of interest. No significant departure from proportionality in hazards over time was found. We analyzed tests for linear trends with linear regression using the ordinal number allocated to each income category. In addition to an unadjusted model (Model 1), we identified potential confounders *a priori* based on the literature review, including age, sex, and residential location in Model 2 to account for sociodemographic characteristics. Model 3 further adjusted for (1) comorbidities that include the presence of chronic kidney disease (CKD), depression, hypertension, and dyslipidemia, and (2) diabetes status and management (fasting glucose concentrations, number of prescriptions for oral anti-diabetic medications per year [<3, ≥3], history of insulin prescription, and T2D duration). We did not include obesity and lifestyle characteristics (smoking status, alcohol consumption, and physical activity) in the main model because of their potential mediating roles in the association between income and CVD risk.^20,35^

We assessed the potential effect modification by age group, sex, health insurance type, and rural/urban residential location using stratified analyses and tests for interaction based on the likelihood ratio test because previous studies showed that the effect of income status on health outcomes might depend on these factors.^36,37^ The following sensitivity analyses were performed: 1) adjustment for potential mediators, including obesity and lifestyle characteristics (Model 4) and 20 initial income quantile at the preceding 4 years (Model 5); 2) using landmark analyses with a 5-year landmark point to estimate the unbiased CVD risk in those who were event-free until 5-year of follow-up and examined the long-term association between income parameters and CVD risk;^38^ and 3) excluding those with a previous history of cancer, which could influence income declines over the years before baseline.^13^ Statistical analyses were performed using SAS version 9.4 (SAS Institute Inc., Cary, NC). The provided *P* values are 2-sided, with the level of significance at 0.05.

## RESULTS

### Baseline characteristics

Baseline characteristics are presented in **table 1** and **Supplementary table 1** by the cumulative number of years in the low- or high-income group and income variability quartile in individuals with T2D. Those with sustained low incomes were more likely to be female, older, never-smokers, non-drinkers, physically inactive, less likely to be self-employed insured, and rural than those who never experienced low income. Individuals with sustained low incomes or high-income variability also were more likely to have central obesity, hypertension, dyslipidemia, CKD, depression, have longer T2D duration (≥5 years), and have used 3 or more oral anti-diabetic or insulin products. Those with sustained high-income status showed opposite patterns compared to those who never experienced high income.

**Table 1.**
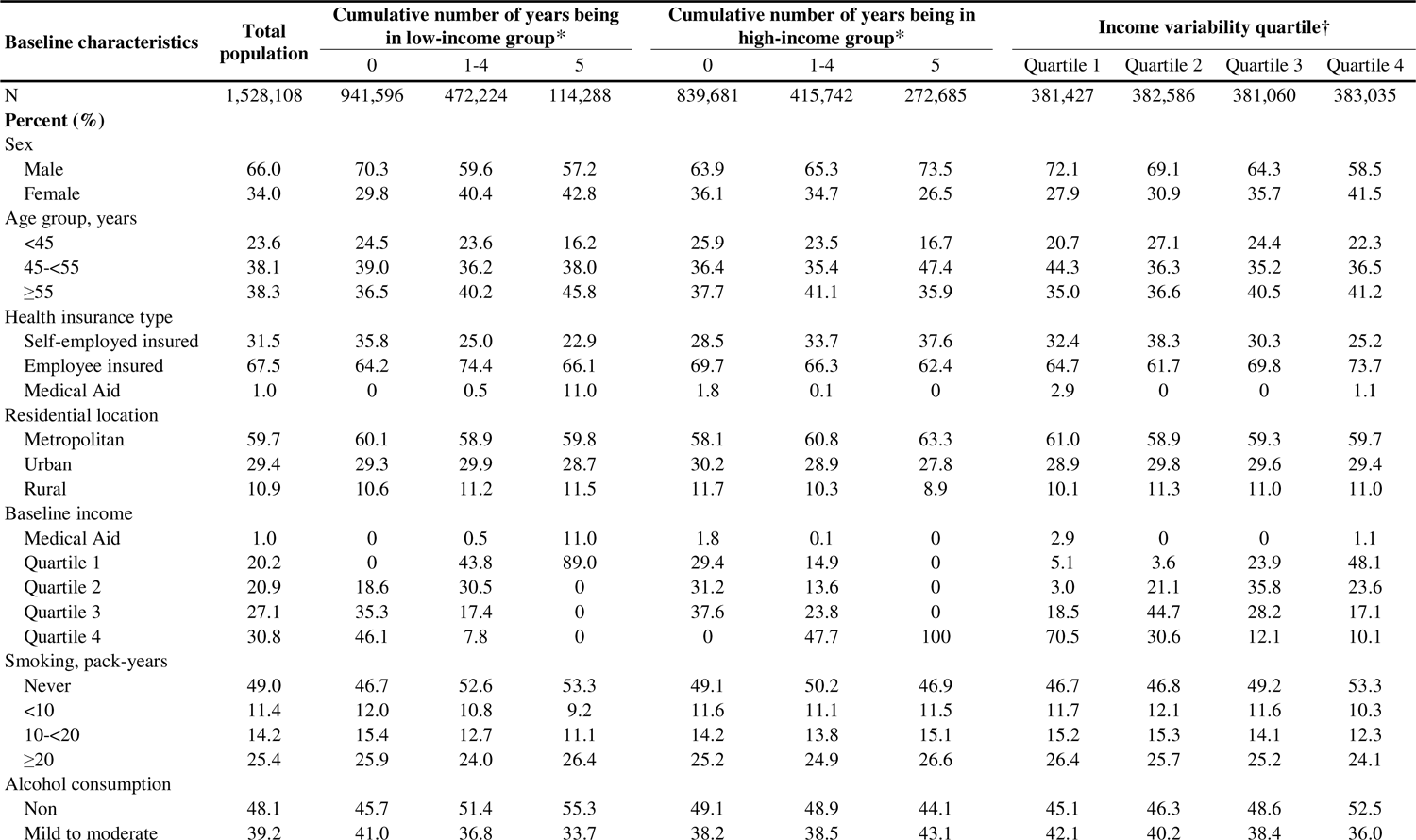
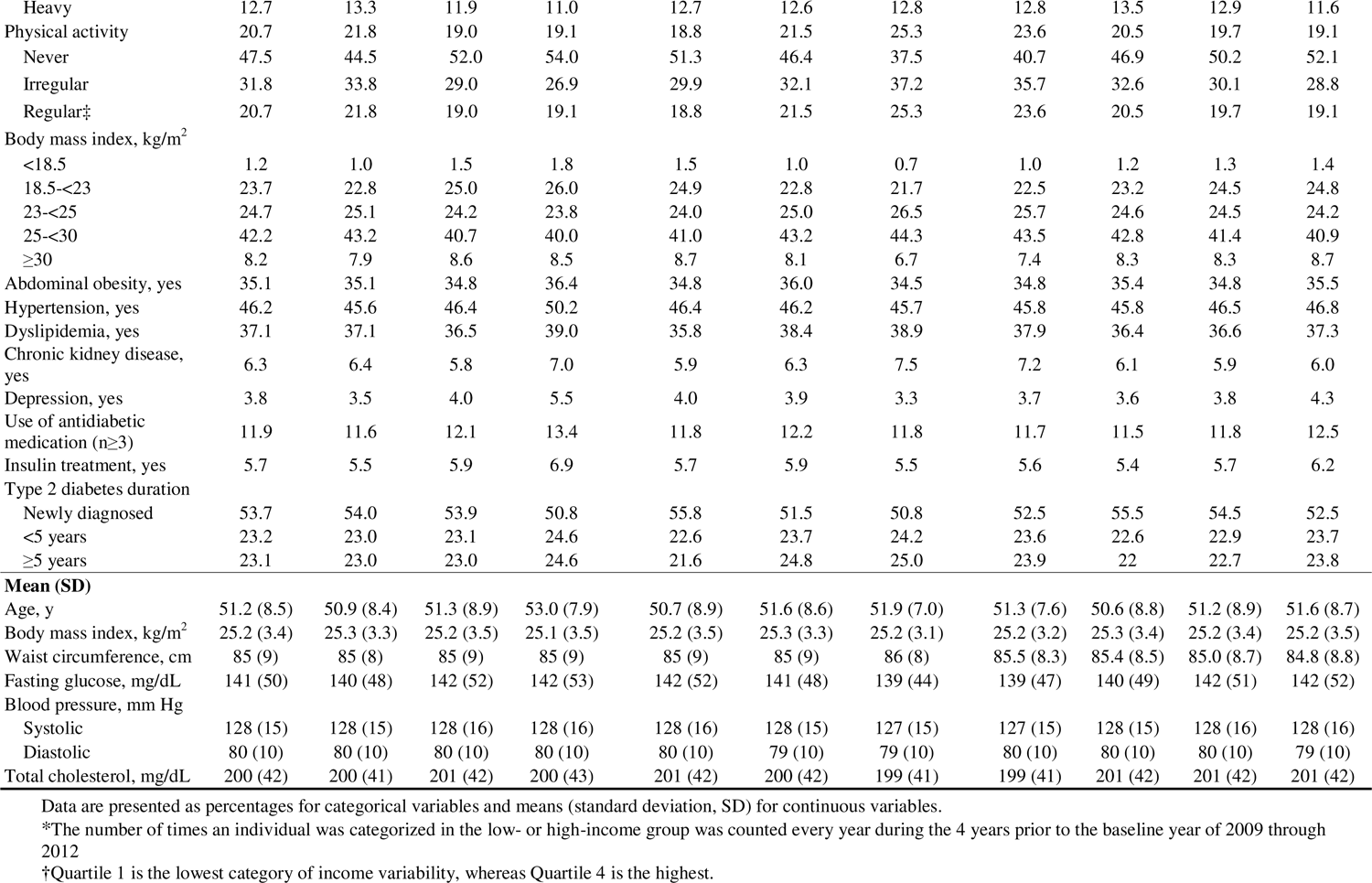
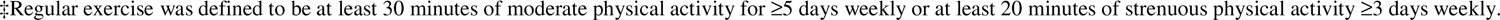
Baseline characteristics by the cumulative number of years being in low- or high-income group and income variability quartile in individuals with type 2 diabetes.

| Baseline characteristics | Total population | Cumulative number of years being in low-income group* |  |  | Cumulative number of years being in high-income group* |  |  | Income variability quartile† |  |  |  |
| --- | --- | --- | --- | --- | --- | --- | --- | --- | --- | --- | --- |
|  |  | 0 | 1-4 | 5 | 0 | 1-4 | 5 | Quartile 1 | Quartile 2 | Quartile 3 | Quartile 4 |
| N | 1,528,108 | 941,596 | 472,224 | 114,288 | 839,681 | 415,742 | 272,685 | 381,427 | 382,586 | 381,060 | 383,035 |
| **Percent (%)** |  |  |  |  |  |  |  |  |  |  |  |
| Sex |  |  |  |  |  |  |  |  |  |  |  |
| Male | 66.0 | 70.3 | 59.6 | 57.2 | 63.9 | 65.3 | 73.5 | 72.1 | 69.1 | 64.3 | 58.5 |
| Female | 34.0 | 29.8 | 40.4 | 42.8 | 36.1 | 34.7 | 26.5 | 27.9 | 30.9 | 35.7 | 41.5 |
| Age group, years |  |  |  |  |  |  |  |  |  |  |  |
| <45 | 23.6 | 24.5 | 23.6 | 16.2 | 25.9 | 23.5 | 16.7 | 20.7 | 27.1 | 24.4 | 22.3 |
| 45-<55 | 38.1 | 39.0 | 36.2 | 38.0 | 36.4 | 35.4 | 47.4 | 44.3 | 36.3 | 35.2 | 36.5 |
| ≥55 | 38.3 | 36.5 | 40.2 | 45.8 | 37.7 | 41.1 | 35.9 | 35.0 | 36.6 | 40.5 | 41.2 |
| Health insurance type |  |  |  |  |  |  |  |  |  |  |  |
| Self-employed insured | 31.5 | 35.8 | 25.0 | 22.9 | 28.5 | 33.7 | 37.6 | 32.4 | 38.3 | 30.3 | 25.2 |
| Employee insured | 67.5 | 64.2 | 74.4 | 66.1 | 69.7 | 66.3 | 62.4 | 64.7 | 61.7 | 69.8 | 73.7 |
| Medical Aid | 1.0 | 0 | 0.5 | 11.0 | 1.8 | 0.1 | 0 | 2.9 | 0 | 0 | 1.1 |
| Residential location |  |  |  |  |  |  |  |  |  |  |  |
| Metropolitan | 59.7 | 60.1 | 58.9 | 59.8 | 58.1 | 60.8 | 63.3 | 61.0 | 58.9 | 59.3 | 59.7 |
| Urban | 29.4 | 29.3 | 29.9 | 28.7 | 30.2 | 28.9 | 27.8 | 28.9 | 29.8 | 29.6 | 29.4 |
| Rural | 10.9 | 10.6 | 11.2 | 11.5 | 11.7 | 10.3 | 8.9 | 10.1 | 11.3 | 11.0 | 11.0 |
| Baseline income |  |  |  |  |  |  |  |  |  |  |  |
| Medical Aid | 1.0 | 0 | 0.5 | 11.0 | 1.8 | 0.1 | 0 | 2.9 | 0 | 0 | 1.1 |
| Quartile 1 | 20.2 | 0 | 43.8 | 89.0 | 29.4 | 14.9 | 0 | 5.1 | 3.6 | 23.9 | 48.1 |
| Quartile 2 | 20.9 | 18.6 | 30.5 | 0 | 31.2 | 13.6 | 0 | 3.0 | 21.1 | 35.8 | 23.6 |
| Quartile 3 | 27.1 | 35.3 | 17.4 | 0 | 37.6 | 23.8 | 0 | 18.5 | 44.7 | 28.2 | 17.1 |
| Quartile 4 | 30.8 | 46.1 | 7.8 | 0 | 0 | 47.7 | 100 | 70.5 | 30.6 | 12.1 | 10.1 |
| Smoking, pack-years |  |  |  |  |  |  |  |  |  |  |  |
| Never | 49.0 | 46.7 | 52.6 | 53.3 | 49.1 | 50.2 | 46.9 | 46.7 | 46.8 | 49.2 | 53.3 |
| <10 | 11.4 | 12.0 | 10.8 | 9.2 | 11.6 | 11.1 | 11.5 | 11.7 | 12.1 | 11.6 | 10.3 |
| 10-<20 | 14.2 | 15.4 | 12.7 | 11.1 | 14.2 | 13.8 | 15.1 | 15.2 | 15.3 | 14.1 | 12.3 |
| ≥20 | 25.4 | 25.9 | 24.0 | 26.4 | 25.2 | 24.9 | 26.6 | 26.4 | 25.7 | 25.2 | 24.1 |
| Alcohol consumption |  |  |  |  |  |  |  |  |  |  |  |
| Non | 48.1 | 45.7 | 51.4 | 55.3 | 49.1 | 48.9 | 44.1 | 45.1 | 46.3 | 48.6 | 52.5 |
| Mild to moderate | 39.2 | 41.0 | 36.8 | 33.7 | 38.2 | 38.5 | 43.1 | 42.1 | 40.2 | 38.4 | 36.0 |
| Heavy | 12.7 | 13.3 | 11.9 | 11.0 | 12.7 | 12.6 | 12.8 | 12.8 | 13.5 | 12.9 | 11.6 |
| Physical activity | 20.7 | 21.8 | 19.0 | 19.1 | 18.8 | 21.5 | 25.3 | 23.6 | 20.5 | 19.7 | 19.1 |
| Never | 47.5 | 44.5 | 52.0 | 54.0 | 51.3 | 46.4 | 37.5 | 40.7 | 46.9 | 50.2 | 52.1 |
| Irregular | 31.8 | 33.8 | 29.0 | 26.9 | 29.9 | 32.1 | 37.2 | 35.7 | 32.6 | 30.1 | 28.8 |
| Regular† | 20.7 | 21.8 | 19.0 | 19.1 | 18.8 | 21.5 | 25.3 | 23.6 | 20.5 | 19.7 | 19.1 |
| Body mass index, kg/m<sup>2</sup> |  |  |  |  |  |  |  |  |  |  |  |
| <18.5 | 1.2 | 1.0 | 1.5 | 1.8 | 1.5 | 1.0 | 0.7 | 1.0 | 1.2 | 1.3 | 1.4 |
| 18.5-<23 | 23.7 | 22.8 | 25.0 | 26.0 | 24.9 | 22.8 | 21.7 | 22.5 | 23.2 | 24.5 | 24.8 |
| 23-<25 | 24.7 | 25.1 | 24.2 | 23.8 | 24.0 | 25.0 | 26.5 | 25.7 | 24.6 | 24.5 | 24.2 |
| 25-<30 | 42.2 | 43.2 | 40.7 | 40.0 | 41.0 | 43.2 | 44.3 | 43.5 | 42.8 | 41.4 | 40.9 |
| ≥30 | 8.2 | 7.9 | 8.6 | 8.5 | 8.7 | 8.1 | 6.7 | 7.4 | 8.3 | 8.3 | 8.7 |
| Abdominal obesity, yes | 35.1 | 35.1 | 34.8 | 36.4 | 34.8 | 36.0 | 34.5 | 34.8 | 35.4 | 34.8 | 35.5 |
| Hypertension, yes | 46.2 | 45.6 | 46.4 | 50.2 | 46.4 | 46.2 | 45.7 | 45.8 | 45.8 | 46.5 | 46.8 |
| Dyslipidemia, yes | 37.1 | 37.1 | 36.5 | 39.0 | 35.8 | 38.4 | 38.9 | 37.9 | 36.4 | 36.6 | 37.3 |
| Chronic kidney disease, yes | 6.3 | 6.4 | 5.8 | 7.0 | 5.9 | 6.3 | 7.5 | 7.2 | 6.1 | 5.9 | 6.0 |
| Depression, yes | 3.8 | 3.5 | 4.0 | 5.5 | 4.0 | 3.9 | 3.3 | 3.7 | 3.6 | 3.8 | 4.3 |
| Use of antidiabetic medication (n≥3) | 11.9 | 11.6 | 12.1 | 13.4 | 11.8 | 12.2 | 11.8 | 11.7 | 11.5 | 11.8 | 12.5 |
| Insulin treatment, yes | 5.7 | 5.5 | 5.9 | 6.9 | 5.7 | 5.9 | 5.5 | 5.6 | 5.4 | 5.7 | 6.2 |
| Type 2 diabetes duration |  |  |  |  |  |  |  |  |  |  |  |
| Newly diagnosed | 53.7 | 54.0 | 53.9 | 50.8 | 55.8 | 51.5 | 50.8 | 52.5 | 55.5 | 54.5 | 52.5 |
| <5 years | 23.2 | 23.0 | 23.1 | 24.6 | 22.6 | 23.7 | 24.2 | 23.6 | 22.6 | 22.9 | 23.7 |
| ≥5 years | 23.1 | 23.0 | 23.0 | 24.6 | 21.6 | 24.8 | 25.0 | 23.9 | 22 | 22.7 | 23.8 |
| **Mean (SD)** |  |  |  |  |  |  |  |  |  |  |  |
| Age, y | 51.2 (8.5) | 50.9 (8.4) | 51.3 (8.9) | 53.0 (7.9) | 50.7 (8.9) | 51.6 (8.6) | 51.9 (7.0) | 51.3 (7.6) | 50.6 (8.8) | 51.2 (8.9) | 51.6 (8.7) |
| Body mass index, kg/m<sup>2</sup> | 25.2 (3.4) | 25.3 (3.3) | 25.2 (3.5) | 25.1 (3.5) | 25.2 (3.5) | 25.3 (3.3) | 25.2 (3.1) | 25.2 (3.2) | 25.3 (3.4) | 25.2 (3.4) | 25.2 (3.5) |
| Waist circumference, cm | 85 (9) | 85 (8) | 85 (9) | 85 (9) | 85 (9) | 85 (9) | 86 (8) | 85.5 (8.3) | 85.4 (8.5) | 85.0 (8.7) | 84.8 (8.8) |
| Fasting glucose, mg/dL | 141 (50) | 140 (48) | 142 (52) | 142 (53) | 142 (52) | 141 (48) | 139 (44) | 139 (47) | 140 (49) | 142 (51) | 142 (52) |
| Blood pressure, mm Hg |  |  |  |  |  |  |  |  |  |  |  |
| Systolic | 128 (15) | 128 (15) | 128 (16) | 128 (16) | 128 (16) | 128 (15) | 127 (15) | 127 (15) | 128 (15) | 128 (16) | 128 (16) |
| Diastolic | 80 (10) | 80 (10) | 80 (10) | 80 (10) | 80 (10) | 79 (10) | 79 (10) | 80 (10) | 80 (10) | 80 (10) | 79 (10) |
| Total cholesterol, mg/dL | 200 (42) | 200 (41) | 201 (42) | 200 (43) | 201 (42) | 200 (42) | 199 (41) | 199 (41) | 201 (42) | 201 (42) | 201 (42) |

Data are presented as percentages for categorical variables and means (standard deviation, SD) for continuous variables.
\*The number of times an individual was categorized in the low- or high-income group was counted every year during the 4 years prior to the baseline year of 2009 through 2012
†Quartile 1 is the lowest category of income variability, whereas Quartile 4 is the highest.
‡Regular exercise was defined to be at least 30 minutes of moderate physical activity for $\geq 5$ days weekly or at least 20 minutes of strenuous physical activity $\geq 3$ days weekly.

Characteristics by income quartiles at baseline are shown in **Supplementary table 2**. Individuals with lower income variability (quartiles 1-2) tended not to experience low income, whereas individuals with higher income variability (quartiles 3-4) tended not to experience high income (**Supplementary table 3**).

### Association of the cumulative number of years in the low- or high-income group and income variability with CVD risk

During follow-up (mean, 7.3 and SD, 1.5 years), 109,319 individuals had a CVD event (7.2%) at least 1 year after baseline. The cumulative incidence of CVD for each income parameter is shown in **fig 2**. **Table 2** shows that individuals with sustained low income over 5 years had the highest CVD risk compared with those who had never experienced low income (Model 3: HR_n=5 years vs. n=0 years_ 1.38, 95% CI 1.35-1.41; *P* for trend <0.0001). Higher-income variability was also associated with increased CVD risk (Model 3: HR_highest vs. lowest quartile_ 1.25, 95% CI 1.22-1.27; *P* for trend<0.0001). The lowest income status (Medical Aid beneficiaries) at baseline exhibited the highest CVD risk compared to those with high income (Model 3: HR 2.95, 95% CI 2.81-3.09; *P* for trend<0.001). In contrast, those with sustained high income had the lowest CVD risk compared to those who never experienced high income (Model 3: HR_n=5 years vs. n=0 years_ 0.71, 95% CI 0.70-0.72; *P* for trend<0.0001). In addition, the strength of associations tended to increase sharply in 5 cumulative very low- or high-income years. When further adjusted for smoking status, alcohol consumption, physical activity, BMI, high waist circumference, and initial income quantile in the preceding 4 years, the associations were attenuated, but the trends were similar (**Supplementary table 4**).

**Figure 2.**
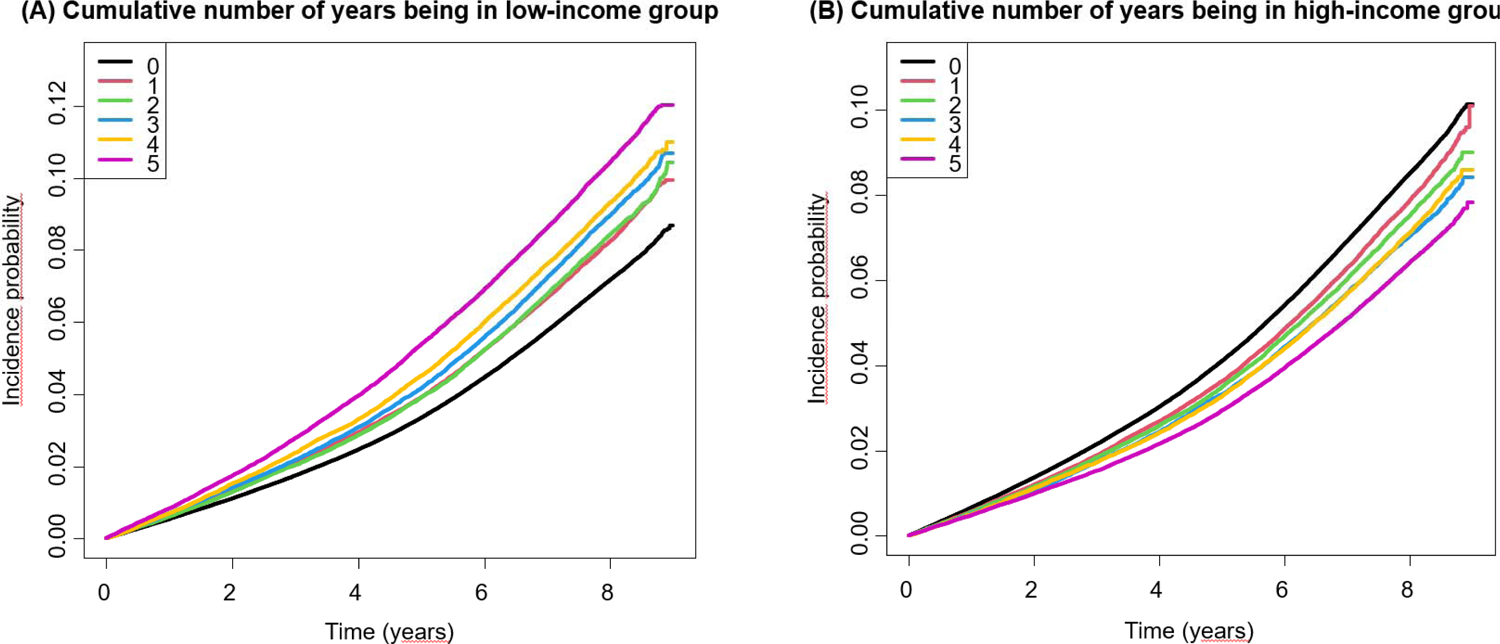
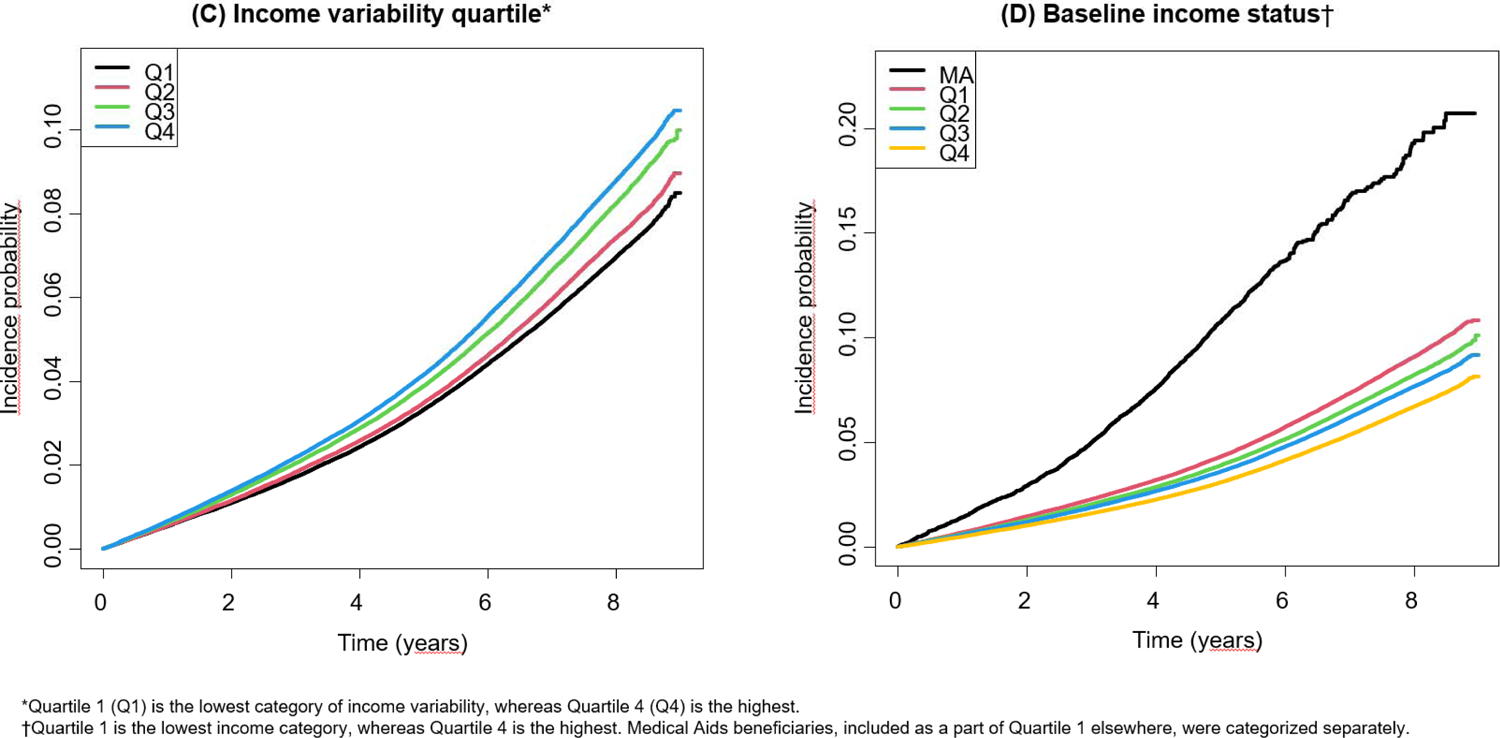
Cumulative incidence function plot for composite cardiovascular events by the cumulative number of years being in (A) low- and (B) high-income group, (C) income variability quartile, and (D) baseline income status Cumulative incidence function plots display composite events of incident fatal and nonfatal cardiovascular diseases by various parameters of income dynamics over the follow-up. A cumulative number of years in the low- or high-income group was counted for 4 years before the baseline year. Income variability was calculated using 20 income quantiles analyzed as a continuous variable to summarize income fluctuations over the same study period.

**Table 2.**
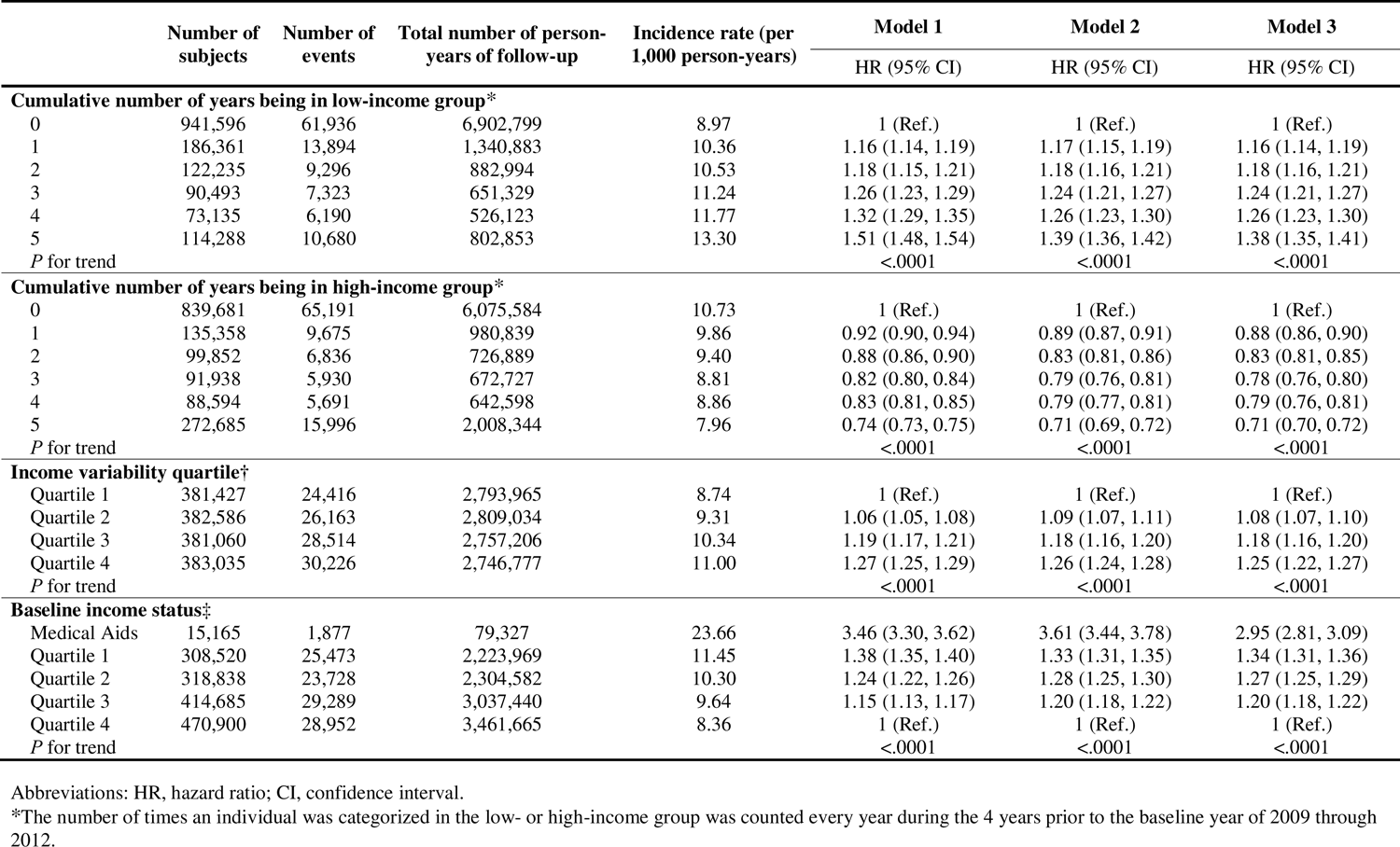
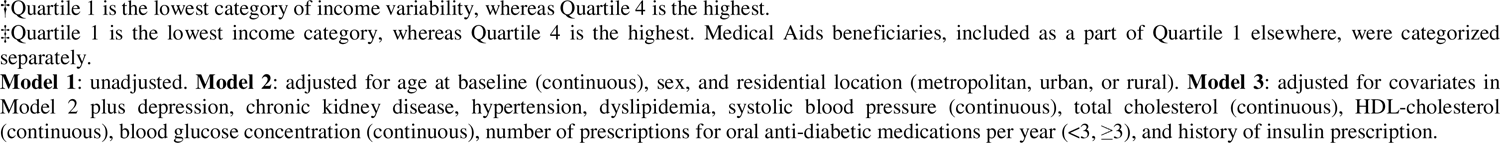
Association between various indicators of income dynamics and risk of composite cardiovascular events in adults with type 2 diabetes.

|  | Number of subjects | Number of events | Total number of person-years of follow-up | Incidence rate (per 1,000 person-years) | Model 1<br>HR (95% CI) | Model 2<br>HR (95% CI) | Model 3<br>HR (95% CI) |
| --- | --- | --- | --- | --- | --- | --- | --- |
| **Cumulative number of years being in low-income group\*** |  |  |  |  |  |  |  |
| 0 | 941,596 | 61,936 | 6,902,799 | 8.97 | 1 (Ref.) | 1 (Ref.) | 1 (Ref.) |
| 1 | 186,361 | 13,894 | 1,340,883 | 10.36 | 1.16 (1.14, 1.19) | 1.17 (1.15, 1.19) | 1.16 (1.14, 1.19) |
| 2 | 122,235 | 9,296 | 882,994 | 10.53 | 1.18 (1.15, 1.21) | 1.18 (1.16, 1.21) | 1.18 (1.16, 1.21) |
| 3 | 90,493 | 7,323 | 651,329 | 11.24 | 1.26 (1.23, 1.29) | 1.24 (1.21, 1.27) | 1.24 (1.21, 1.27) |
| 4 | 73,135 | 6,190 | 526,123 | 11.77 | 1.32 (1.29, 1.35) | 1.26 (1.23, 1.30) | 1.26 (1.23, 1.30) |
| 5 | 114,288 | 10,680 | 802,853 | 13.30 | 1.51 (1.48, 1.54) | 1.39 (1.36, 1.42) | 1.38 (1.35, 1.41) |
| *P* for trend |  |  |  |  | <.0001 | <.0001 | <.0001 |
| **Cumulative number of years being in high-income group\*** |  |  |  |  |  |  |  |
| 0 | 839,681 | 65,191 | 6,075,584 | 10.73 | 1 (Ref.) | 1 (Ref.) | 1 (Ref.) |
| 1 | 135,358 | 9,675 | 980,839 | 9.86 | 0.92 (0.90, 0.94) | 0.89 (0.87, 0.91) | 0.88 (0.86, 0.90) |
| 2 | 99,852 | 6,836 | 726,889 | 9.40 | 0.88 (0.86, 0.90) | 0.83 (0.81, 0.86) | 0.83 (0.81, 0.85) |
| 3 | 91,938 | 5,930 | 672,727 | 8.81 | 0.82 (0.80, 0.84) | 0.79 (0.76, 0.81) | 0.78 (0.76, 0.80) |
| 4 | 88,594 | 5,691 | 642,598 | 8.86 | 0.83 (0.81, 0.85) | 0.79 (0.77, 0.81) | 0.79 (0.76, 0.81) |
| 5 | 272,685 | 15,996 | 2,008,344 | 7.96 | 0.74 (0.73, 0.75) | 0.71 (0.69, 0.72) | 0.71 (0.70, 0.72) |
| *P* for trend |  |  |  |  | <.0001 | <.0001 | <.0001 |
| **Income variability quartile†** |  |  |  |  |  |  |  |
| Quartile 1 | 381,427 | 24,416 | 2,793,965 | 8.74 | 1 (Ref.) | 1 (Ref.) | 1 (Ref.) |
| Quartile 2 | 382,586 | 26,163 | 2,809,034 | 9.31 | 1.06 (1.05, 1.08) | 1.09 (1.07, 1.11) | 1.08 (1.07, 1.10) |
| Quartile 3 | 381,060 | 28,514 | 2,757,206 | 10.34 | 1.19 (1.17, 1.21) | 1.18 (1.16, 1.20) | 1.18 (1.16, 1.20) |
| Quartile 4 | 383,035 | 30,226 | 2,746,777 | 11.00 | 1.27 (1.25, 1.29) | 1.26 (1.24, 1.28) | 1.25 (1.22, 1.27) |
| *P* for trend |  |  |  |  | <.0001 | <.0001 | <.0001 |
| **Baseline income status‡** |  |  |  |  |  |  |  |
| Medical Aids | 15,165 | 1,877 | 79,327 | 23.66 | 3.46 (3.30, 3.62) | 3.61 (3.44, 3.78) | 2.95 (2.81, 3.09) |
| Quartile 1 | 308,520 | 25,473 | 2,223,969 | 11.45 | 1.38 (1.35, 1.40) | 1.33 (1.31, 1.35) | 1.34 (1.31, 1.36) |
| Quartile 2 | 318,838 | 23,728 | 2,304,582 | 10.30 | 1.24 (1.22, 1.26) | 1.28 (1.25, 1.30) | 1.27 (1.25, 1.29) |
| Quartile 3 | 414,685 | 29,289 | 3,037,440 | 9.64 | 1.15 (1.13, 1.17) | 1.20 (1.18, 1.22) | 1.20 (1.18, 1.22) |
| Quartile 4 | 470,900 | 28,952 | 3,461,665 | 8.36 | 1 (Ref.) | 1 (Ref.) | 1 (Ref.) |
| *P* for trend |  |  |  |  | <.0001 | <.0001 | <.0001 |

Abbreviations: HR, hazard ratio; CI, confidence interval.
\*The number of times an individual was categorized in the low- or high-income group was counted every year during the 4 years prior to the baseline year of 2009 through 2012.
†Quartile 1 is the lowest category of income variability, whereas Quartile 4 is the highest.
‡Quartile 1 is the lowest income category, whereas Quartile 4 is the highest. Medical Aids beneficiaries, included as a part of Quartile 1 elsewhere, were categorized separately.
**Model 1:** unadjusted. **Model 2:** adjusted for age at baseline (continuous), sex, and residential location (metropolitan, urban, or rural). **Model 3:** adjusted for covariates in Model 2 plus depression, chronic kidney disease, hypertension, dyslipidemia, systolic blood pressure (continuous), total cholesterol (continuous), HDL-cholesterol (continuous), blood glucose concentration (continuous), number of prescriptions for oral anti-diabetic medications per year ( $<3$ , $\geq 3$ ), and history of insulin prescription.

Associations between various income parameters and individual CV event types, including MI, stroke, HF, and CV mortality, were generally similar to those of the composite CVD events measure; however, CV mortality showed the strongest associations (Supplementary **table 5)**.

### Association of income changes between the 2 time points (4 years ago vs. baseline) with CVD risk

Associations of changes in income status between the first assessment (4 years before baseline) and the last (baseline) with CVD risk are shown in **fig 3** and **Supplementary table 6**. When we compared each income quartile group between the time points, CVD risk linearly decreased with greater income rise and increased with greater income decline compared to persistent income status. Meanwhile, regardless of initial income status, those who experienced income decline to the level of Medical Aid beneficiaries had over 2-fold increased CVD risk. Similar associations were also observed in individual CVD outcomes (**Supplementary figure 1**).

**Figure 3.**
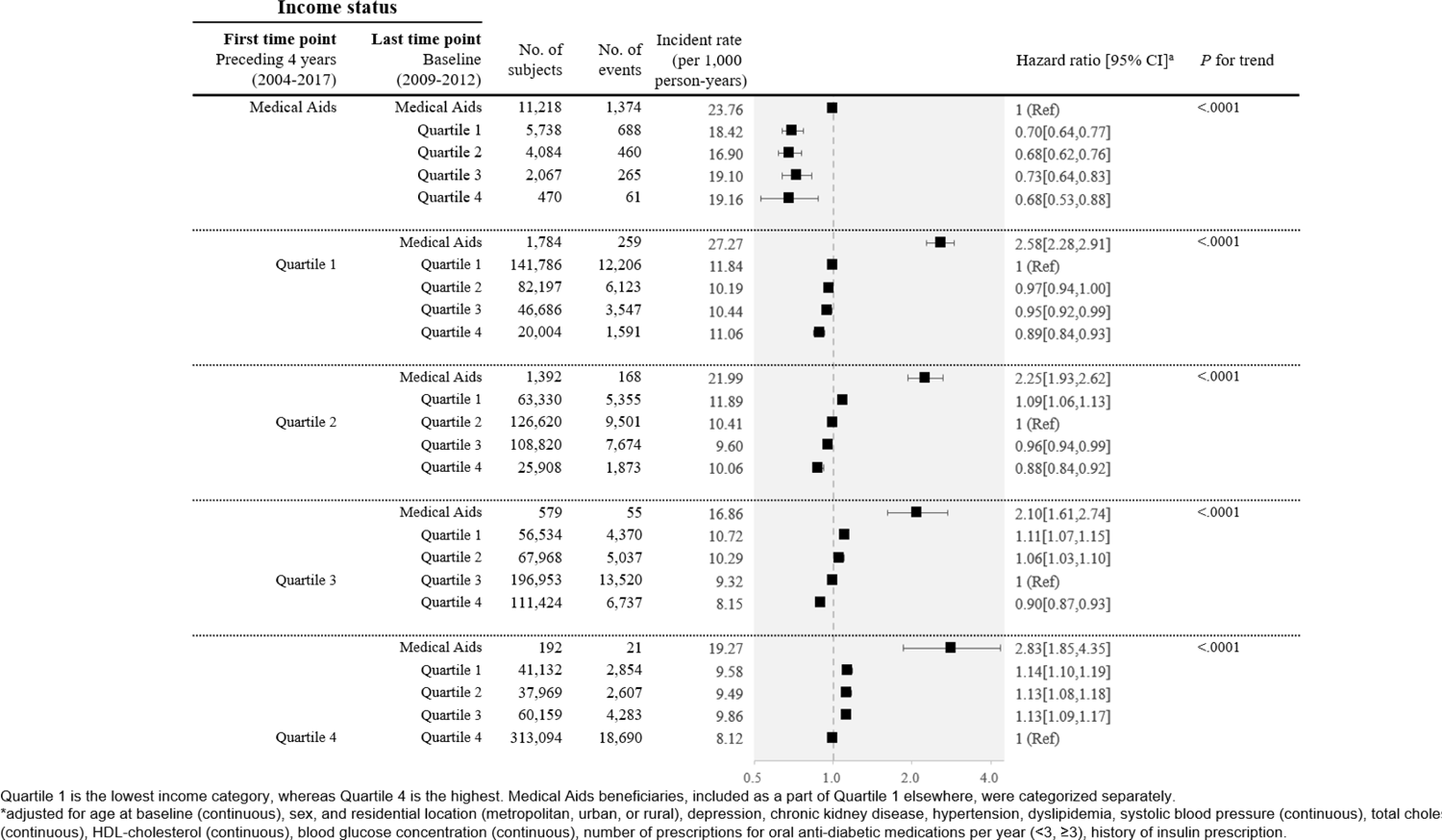
Change of income status between the 2 time points (4 years ago vs. baseline) and the corresponding risk of composite cardiovascular events Hazard ratios and 95% confidence intervals are shown in the association of changes in income status between the first assessment (4 years before baseline) and the last (baseline) with composite incident events of fatal and nonfatal cardiovascular disease. In each income group, individuals who experienced income rise or decline are compared with those with a persistent income status between the 2 time points.

### Stratified and Sensitivity analyses

In stratified analyses by potential effect modifiers of the association between income dynamics and CVD risk, the association of the number of years being in the low-income group and high-income variability quartile with CVD risk was stronger for those who were younger (<45 years), males, self-employed insured, and non-rural residents (all *P* for interaction<0.05) (**Supplementary figure 2**). The results did not substantially differ after excluding those with a prior history of cancer (**Supplementary table 7**) and when our analysis was limited to those who were event-free until the 5-year follow-up (**Supplementary table 8**).

## DISCUSSION

In our nationwide cohort study of over 1.5 million non-elderly adults with T2D, higher income variability and sustained low income over 5 years were associated with increased CVD risk. Individuals who consecutively had low income over 5 years had 38% increased CVD risk, compared with those who never experienced low income after adjusting for multiple sociodemographic and CV risk factors, and T2D management. In contrast, individuals who consecutively had high income over 5 years had 29% decreased CVD risk, compared with those who never experienced high income. Individuals who experienced an income decline to the lowest income level (i.e., Medical Aid beneficiaries) had over a 2-fold increased CVD risk compared to those with persistent income status, regardless of their initial income status. All these associations remained robust after adjusting for potential mediators (e.g., obesity and lifestyle factors).

Prior studies examining income and CVD risk in individuals with T2D were based on a one-time measurement of income which ignores the impact of income dynamics over time.^9,11,18,21,22^ Only 3 recent studies reported individual-level income declines^13,26^ and high-income volatility^25^ increase CVD risk among older,^26^ middle-aged,^13^ and young^25^ adults in the general US population. Findings from the prior studies may not apply to ours because individuals with T2D are at high risk for CVD,^39^ and SES and T2D disease status interact with each other to affect CVD risk.^20^ In addition, previous studies evaluated income changes over more than 15 years, which could be more influenced by macroeconomic factors,^25,26^ compared to our study with a short-term income variability over 5 years and use of relative income change.

Individuals with high-income variability had increased CVD risk, and they had low baseline income and were less likely to experience high income over the pre-baseline period. Income variability accompanies income uncertainty or rapid and unpredictable income changes over time, which may result in additional vulnerability, especially in low-income households.^23^ In addition, individuals who experienced an income decline to the lowest income level (Medical Aid beneficiaries) had much greater CVD risk than those with other income declines, regardless of their initial income status. The Medical Aid program was designed to support the medical needs of poor people in South Korea.^32^ Medical Aid beneficiaries pay much smaller out-of-pocket payments for outpatient services and are exempt from hospitalization fees.^40^ As a result, they tend to use more healthcare services (outpatient visits and hospitalization), and their medical costs per person are higher than for NHI-covered individuals.^41,42^ Poor health behaviors, and irregular and suboptimal assessment for comorbidities could still explain their health disparities.^20^ Interventions to address CV risk factors may not be equally distributed across SES groups, and socioeconomically deprived individuals could receive paradoxically fewer benefits.^43^ Thus, an integrative, collaborative approach may be required to reduce clinical and public health disparities among those with low income.^2,43^

Any low-income experience within 4 years before baseline was associated with increased CVD risk. A dose-response relationship was also observed in which CVD risk increased linearly with the years of being in low income. The longer income is unstable or remains low, the more likely the development of unhealthy lifestyles, food insecurity, and comorbidities.^44^ Consequently, individuals exposed or worsened to low-income status may be more vulnerable to CVD because those with low SES had poor access to optimal medication use and adherence, achieving suboptimal control targets for CV risk factors.^45,46^ Another study demonstrated that multiple job losses were associated with increased MI risk in a dose-response manner.^47^ Our previous study also showed a similar pattern in which the risk of all-cause mortality increased incrementally with an increasing number of low-income status.^33^

In contrast, exposure to high-income status over the 4 years before baseline was associated with decreased CVD risk, with the lowest risk when sustained high income was maintained. Also, any increase in income status over these 5 years was associated with decreased CVD risk. Other investigators showed a long-term CV risk reduction associated with a 50% income rise over 6 years in the general population.^13^ Higher incomes increase healthcare quality and utilization.^48^ Thus, income increases and maintaining a stable income may be linked to favorable CVD outcomes.

Ours is the first study to examine the longitudinal association between income dynamics and CVD risk in non-elderly adults with T2D, considering important confounders, mediators, and effect modifiers, particularly in a setting of universal health coverage where income status has little effect on access to medical services.^32^ Thus, our findings might not be affected by healthcare accessibility.^41^ Additionally, laboratory findings or anthropometric parameters were collected systematically through a validated national standard protocol.^49^

Limitations include our lack of access to other covariates, including food security, medication adherence, educational attainment, or laboratory results about achieving treatment goals. Data about the number of household members contributing to or supported by household income was also unavailable. Finally, the results may not apply to other populations with different ethnic, socioeconomic, and cultural backgrounds.

In conclusion, higher income variability, an income decline, and sustained low-income status were associated with increased CVD risk, whereas sustained high-income status and income increase were associated with decreased CVD risk in over 1.5 million non-elderly Korean adults with T2D. Our findings provide evidence for the role of income dynamics in CVD risk in individuals with T2D and underscore the need for increased public policy awareness of the impact of income dynamics on CVD risk in non-elderly adults with T2D.

## ETHICAL APPROVAL

The study protocol was approved by the institutional review boards of Soongsil University. The study makes use of routinely collected data that was de- personalized before release. Informed consent from individual participants was therefore not required.

## Data Availability

All data produced in the present work are contained in the manuscript.

## ACKNOWLEDGMENT

Christopher B McLeod, PhD (Partnership for Work, Health and Safety, School of Population and Public Health, University of British Columbia, Vancouver, British Columbia, Canada) provided helpful comments.

