## supplements for "Longitudinal associations of sustained low or high income and income variability with incident cardiovascular disease in individuals with type 2 diabetes: a retrospective population-based cohort study"

**eMethods.** Supplemental methods

**Supplementary table 1**. Baseline characteristics by cumulative number of years (0, 1, 2, 3, 4, and 5) being in low- or high-income group in individuals with type 2 diabetes

**Supplementary table 2**. Baseline characteristics by baseline income status in individuals with type 2 diabetes

**Supplementary table 3**. Distribution of cumulative number of years being in low- or high-income group according to income variability quartile

**Supplementary table 4**. Association between various indicators of income dynamics and risk of composite cardiovascular events in individuals with type 2 diabetes, with further adjustment for the potential mediators such as obesity and lifestyle factors, as well as initial income quantiles in the preceding 4 years

**Supplementary table 5**. Association between various indicators of income dynamics and the corresponding risk of individual cardiovascular events in adults with type 2 diabetes

**Supplementary table 6**. Changes in income status between the two time points (4 years ago vs. baseline) and the corresponding risk of composite cardiovascular events in individuals with type 2 diabetes, with further adjustment for the potential mediators such as obesity and lifestyle factors, as well as initial income quantiles in the preceding 4 years

**Supplementary table 7**. Association between various indicators of income dynamics and risk of composite cardiovascular events in individuals with type 2 diabetes, after excluding those who had a prior cancer history

**Supplementary table 8**. 5-year landmark analyses of the risk of composite cardiovascular events according to the cumulative number of years being in low- or high-income group, income variability quartile, and baseline income status

**Supplementary figure 1**. Change of income status between the two-time points (4 years ago vs. baseline) and the corresponding risk of individual cardiovascular events in individuals with type 2 diabetes: (A) myocardial infarction, (B) ischemic stroke, (C) heart failure, and (D) cardiovascular mortality

**Supplementary figure 2**. Association between various indicators of income dynamics and risk of composite cardiovascular events in individuals with type 2 diabetes stratified by selected factors

**eMethods**

Information on lifestyle-related factors was obtained using self-administered questionnaires. The cumulative lifetime smoking exposure was calculated as the pack-year by multiplying the average daily number of cigarettes smoked (pack) by the total duration of smoking (years). Alcohol consumption was also categorized into non-drinking, mild to moderate drinking (<30 g/day), and heavy drinking (≥30 g/day).^1^ For physical activity, regular exercise was defined as at least 30 minutes of moderate physical activity for ≥5 days weekly or at least 20 minutes of strenuous physical activity ≥3 days weekly.^2^

The anthropometric and clinical characteristics were assessed by trained personnel during the health screening examination. Body mass index (BMI) was calculated by weight in kilograms divided by the square of height in meters (kg/m^2^). Abdominal obesity is a waist circumference ≥90 cm for men and ≥85 cm for women, according to Korean population standards and guidelines.^3^ Systolic and diastolic blood pressures were measured after at least 5 minutes of rest with the patient in a sitting position. Blood samples were collected after overnight fasting, and glucose and creatinine serum levels were measured. Quality control procedures for laboratory tests were performed based on the Korean Association of Laboratory Quality Control guidelines.^4^

The presence of hypertension was defined as having at least one prescription of antihypertensive medications under ICD-10 codes I10–I15 per year or systolic/diastolic blood pressure ≥140/90 mmHg.^5^ Dyslipidemia was defined as a total cholesterol level of ≥ 240 mg/dL or taking lipid-lowering drugs.^6^ Chronic kidney disease was defined as an estimated glomerular filtration rate <60 mL/min.^7^ Other clinical comorbidities were defined using the ICD-10 codes and the prescription lists in the NHIS database. Depression was defined to have ICD-10 codes corresponding to F22-F23. Newly diagnosed diabetes was defined as having no history of claims for the ICD-10 code of diabetes (E11-E14) or anti-diabetic medication before enrollment and FPG ≥126 mg/dL at enrollment.^8^

**Supplementary table 1. Baseline characteristics by cumulative number of years (0, 1, 2, 3, 4, and 5) being in low- or high-income group in individuals with type 2 diabetes**

|  | | **Cumulative number of years being in low-income group*** | | | | | |  | **Cumulative number of years being in high-income group*** | | | | | |
| --- | --- | --- | --- | --- | --- | --- | --- | --- | --- | --- | --- | --- | --- | --- |
|  | | 0 | 1 | 2 | 3 | 4 | 5 |  | 0 | 1 | 2 | 3 | 4 | 5 |
| N | | 941,596 | 186,361 | 122,235 | 90,493 | 73,135 | 114,288 |  | 839,681 | 135,358 | 99,852 | 91,938 | 88,594 | 272,685 |
| **Percent (%)** | |  |  |  |  |  |  |  |  |  |  |  |  |  |
| Sex | |  |  |  |  |  |  |  |  |  |  |  |  |  |
|  | Male | 70.3 | 61.8 | 59.1 | 57.6 | 57.1 | 57.2 |  | 63.9 | 64.8 | 64.9 | 65.8 | 65.8 | 73.5 |
|  | Female | 29.8 | 38.2 | 40.9 | 42.4 | 42.9 | 42.8 |  | 36.1 | 35.2 | 35.1 | 34.3 | 34.2 | 26.5 |
| Age group, years | |  |  |  |  |  |  |  |  |  |  |  |  |  |
|  | <45 | 24.5 | 25.1 | 24.1 | 22.6 | 20.3 | 16.2 |  | 25.9 | 24.8 | 23.3 | 23.8 | 21.7 | 16.7 |
|  | 45-<55 | 39.0 | 35.7 | 36.3 | 36.5 | 36.7 | 38.0 |  | 36.4 | 34.1 | 35.1 | 35.4 | 37.6 | 47.4 |
|  | ≥55 | 36.5 | 39.2 | 39.6 | 40.9 | 43.0 | 45.8 |  | 37.7 | 41.2 | 41.7 | 40.8 | 40.6 | 35.9 |
| Health insurance type | |  |  |  |  |  |  |  |  |  |  |  |  |  |
|  | Self-employed insured | 35.8 | 26.3 | 25.1 | 24.1 | 22.9 | 22.9 |  | 28.5 | 30.3 | 33.2 | 35.0 | 38.1 | 37.6 |
|  | Employee insured | 64.2 | 73.5 | 74.6 | 75.0 | 75.6 | 66.1 |  | 69.7 | 69.6 | 66.7 | 65.0 | 61.8 | 62.4 |
|  | Medical Aid Beneficiaries | NA | 0.2 | 0.3 | 0.9 | 1.5 | 11.0 |  | 1.8 | 0.1 | 0.1 | 0.0 | 0.0 | 0.0 |
| Residential location | |  |  |  |  |  |  |  |  |  |  |  |  |  |
|  | Metropolitan | 60.1 | 58.9 | 58.8 | 58.8 | 59.6 | 59.8 |  | 58.1 | 60.4 | 60.8 | 60.7 | 61.3 | 63.3 |
|  | Urban | 29.3 | 29.9 | 30.0 | 30.0 | 29.4 | 28.7 |  | 30.2 | 29.3 | 29.0 | 28.8 | 28.4 | 27.8 |
|  | Rural | 10.6 | 11.2 | 11.3 | 11.3 | 11.0 | 11.5 |  | 11.7 | 10.3 | 10.2 | 10.5 | 10.3 | 8.9 |
| Smoking, pack-years | |  |  |  |  |  |  |  |  |  |  |  |  |  |
|  | Never | 46.7 | 51.1 | 52.8 | 53.9 | 54.3 | 53.3 |  | 49.1 | 49.7 | 50.4 | 50.2 | 50.6 | 46.9 |
|  | <10 | 12.0 | 11.3 | 11.1 | 10.2 | 9.5 | 9.2 |  | 11.6 | 11.2 | 11.1 | 11.2 | 10.6 | 11.5 |
|  | 10-<20 | 15.4 | 13.6 | 12.6 | 12.0 | 11.7 | 11.1 |  | 14.2 | 14.1 | 13.7 | 13.8 | 13.7 | 15.1 |
|  | ≥20 | 25.9 | 24.0 | 23.5 | 23.9 | 24.5 | 26.4 |  | 25.2 | 25.0 | 24.8 | 24.8 | 25.1 | 26.6 |
| Alcohol consumption | |  |  |  |  |  |  |  |  |  |  |  |  |  |
|  | Non | 45.7 | 50.1 | 51.3 | 52.4 | 53.5 | 55.3 |  | 49.1 | 49.0 | 49.2 | 48.8 | 48.7 | 44.1 |
|  | Mild to moderate | 41.0 | 37.6 | 36.7 | 36.0 | 35.6 | 33.7 |  | 38.2 | 38.3 | 38.2 | 38.7 | 38.9 | 43.1 |
|  | Heavy | 13.3 | 12.3 | 12.0 | 11.6 | 11.0 | 11.0 |  | 12.7 | 12.8 | 12.6 | 12.6 | 12.4 | 12.8 |
| Physical activity | |  |  |  |  |  |  |  |  |  |  |  |  |  |
|  | Never | 44.5 | 51.2 | 51.9 | 52.8 | 52.9 | 54.0 |  | 51.3 | 48.4 | 47.0 | 45.0 | 44.0 | 37.5 |
|  | Irregular | 33.8 | 29.6 | 29.3 | 28.4 | 27.9 | 26.9 |  | 29.9 | 31.2 | 31.9 | 32.8 | 33.0 | 37.2 |
|  | Regular† | 21.8 | 19.2 | 18.8 | 18.7 | 19.2 | 19.1 |  | 18.8 | 20.4 | 21.1 | 22.1 | 23.0 | 25.3 |
| Body mass index, kg/m^2^ | |  |  |  |  |  |  |  |  |  |  |  |  |  |
|  | <18.5 | 1.0 | 1.4 | 1.5 | 1.5 | 1.6 | 1.8 |  | 1.5 | 1.2 | 1.0 | 1.0 | 0.9 | 0.7 |
|  | 18.5-<23 | 22.8 | 24.6 | 24.9 | 25.6 | 25.6 | 26.0 |  | 24.9 | 23.2 | 22.8 | 22.6 | 22.3 | 21.7 |
|  | 23-<25 | 25.1 | 24.4 | 24.2 | 24.0 | 24.0 | 23.8 |  | 24.0 | 24.6 | 24.9 | 25.0 | 25.6 | 26.5 |
|  | 25-<30 | 43.2 | 40.9 | 40.8 | 40.4 | 40.4 | 40.0 |  | 41.0 | 42.7 | 43.2 | 43.7 | 43.4 | 44.3 |
|  | ≥30 | 7.9 | 8.7 | 8.7 | 8.5 | 8.4 | 8.5 |  | 8.7 | 8.4 | 8.1 | 7.8 | 7.8 | 6.7 |
| Abdominal obesity, yes | | 35.1 | 34.8 | 34.8 | 34.6 | 35.3 | 36.4 |  | 34.8 | 36.1 | 36.2 | 36.1 | 35.8 | 34.5 |
| Hypertension, yes | | 45.6 | 45.8 | 46.0 | 46.7 | 48.0 | 50.2 |  | 46.4 | 46.3 | 46.3 | 46.1 | 46.0 | 45.7 |
| Dyslipidemia, yes | | 37.1 | 36.4 | 36.5 | 36.3 | 37.1 | 39.0 |  | 35.8 | 37.9 | 38.6 | 38.3 | 39.1 | 38.9 |
| Chronic kidney disease, yes | | 6.4 | 5.7 | 5.7 | 5.9 | 6.2 | 7.0 |  | 5.9 | 6.1 | 6.3 | 6.5 | 6.5 | 7.5 |
| Depression, yes | | 3.5 | 3.9 | 4.0 | 4.1 | 4.3 | 5.5 |  | 4.0 | 3.9 | 4.0 | 3.9 | 3.9 | 3.3 |
| Use of antidiabetic medication (n≥3) | | 11.6 | 11.9 | 12.0 | 12.4 | 12.4 | 13.4 |  | 11.8 | 12.1 | 12.2 | 12.3 | 12.4 | 11.8 |
| Insulin treatment, yes | | 5.5 | 5.8 | 5.8 | 6.0 | 6.2 | 6.9 |  | 5.7 | 6.0 | 5.8 | 5.9 | 5.9 | 5.5 |
| Type 2 diabetes duration | |  |  |  |  |  |  |  |  |  |  |  |  |  |
| Newly diagnosed | | 54.0 | 54.2 | 54.2 | 53.8 | 53.0 | 50.8 |  | 55.8 | 52.7 | 51.3 | 51.5 | 50.1 | 50.8 |
|  | <5 years | 23.0 | 23.0 | 22.9 | 23.3 | 23.8 | 24.6 |  | 22.6 | 23.1 | 24.0 | 23.6 | 24.3 | 24.2 |
|  | ≥5 years | 23.0 | 22.9 | 22.9 | 23.0 | 23.2 | 24.6 |  | 21.6 | 24.2 | 24.8 | 24.9 | 25.6 | 25.0 |
| **Mean (SD)** | |  |  |  |  |  |  |  |  |  |  |  |  |  |
| Age, y | | 50.9 (8.4) | 50.9 (9.0) | 51.1 (8.9) | 51.5 (8.7) | 52.1 (8.5) | 53.0 (7.9) |  | 50.7 (8.9) | 51.3 (9.0) | 51.6 (8.7) | 51.6 (8.6) | 51.9 (8.1) | 51.9 (7.0) |
| Body mass index, kg/m^2^ | | 25.3 (3.3) | 25.2 (3.5) | 25.2 (3.5) | 25.1 (3.5) | 25.1 (3.5) | 25.1 (3.5) |  | 25.2 (3.5) | 25.3 (3.4) | 25.3 (3.3) | 25.3 (3.3) | 25.3 (3.2) | 25.2 (3.1) |
| Waist circumference, cm | | 85.4 (8.4) | 84.9 (8.8) | 84.7 (8.8) | 84.5 (8.9) | 84.6 (8.9) | 84.8 (9.0) |  | 84.9 (8.8) | 85.3 (8.6) | 85.3 (8.5) | 85.4 (8.5) | 85.4 (8.4) | 85.6 (8.1) |
| Fasting glucose, mg/dL | | 140 (48) | 142 (51) | 142 (51) | 142 (52) | 142 (52) | 142 (53) |  | 142 (52) | 142 (50) | 141 (49) | 140 (47) | 140 (47) | 139 (44) |
| Blood pressure, mm Hg | |  |  |  |  |  |  |  |  |  |  |  |  |  |
|  | Systolic | 128 (15) | 128 (16) | 128 (16) | 128 (16) | 128 (16) | 128 (16) |  | 128 (16) | 128 (15) | 128 (15) | 127 (15) | 127 (15) | 127 (15) |
|  | Diastolic | 80 (10) | 80 (10) | 79 (10) | 79 (10) | 80 (11) | 80 (10) |  | 80 (10) | 79 (10) | 79 (10) | 79 (10) | 79 (10) | 79 (10) |
| Total cholesterol, mg/dL | | 200 (41) | 201 (42) | 201 (42) | 201 (42) | 201 (42) | 200 (43) |  | 201 (42) | 200 (42) | 200 (42) | 200 (41) | 199 (41) | 199 (41) |

Data are presented as percentages for categorical variables and means (standard deviation) for continuous variables.

*The number of times an individual was categorized in the low- or high-income group was counted every year during the 5 years prior to the baseline year of 2009 through 2012

†Regular exercise was defined to be at least 30 minutes of moderate physical activity for ≥5 days weekly or at least 20 minutes of strenuous physical activity ≥3 days weekly

**Supplementary table 2. Baseline characteristics by baseline income status in individuals with type 2 diabetes**

|  | | **Baseline income status*** | | | | |
| --- | --- | --- | --- | --- | --- | --- |
|  | | Medical Aids Beneficiaries | Quartile 1 | Quartile 2 | Quartile 3 | Quartile 4 |
| N | | 15,165 | 308,520 | 318,838 | 414,685 | 470,900 |
| **Percent (%)** | |  |  |  |  |  |
| Sex | |  |  |  |  |  |
|  | Male | 50.8 | 58.3 | 65.0 | 68.0 | 70.4 |
|  | Female | 49.2 | 41.7 | 35.0 | 32.0 | 29.6 |
| Age group, years | |  |  |  |  |  |
|  | <45 | 21.6 | 19.4 | 25.5 | 28.1 | 21.3 |
|  | 45-<55 | 39.5 | 37.6 | 37.3 | 34.8 | 41.8 |
|  | ≥55 | 38.9 | 43.0 | 37.2 | 37.2 | 37.0 |
| Health insurance type | |  |  |  |  |  |
|  | Self-employed insured | NA | 18.7 | 30.1 | 34.5 | 39.3 |
|  | Employee insured | 0.0 | 81.3 | 70.0 | 65.5 | 60.7 |
|  | Medical Aid Beneficiaries | 100.0 | NA | NA | NA | NA |
| Residential location | |  |  |  |  |  |
|  | Metropolitan | 53.5 | 60.4 | 58.1 | 58.2 | 62.0 |
|  | Urban | 31.1 | 28.9 | 30.1 | 30.2 | 28.5 |
|  | Rural | 15.4 | 10.7 | 11.8 | 11.6 | 9.5 |
| Smoking, pack-years | |  |  |  |  |  |
|  | Never | 55.0 | 53.5 | 48.3 | 47.0 | 48.0 |
|  | <10 | 10.8 | 9.4 | 12.0 | 12.4 | 11.5 |
|  | 10-<20 | 10.4 | 11.7 | 14.4 | 15.5 | 14.9 |
|  | ≥20 | 23.9 | 25.4 | 25.3 | 25.1 | 25.6 |
| Alcohol consumption | |  |  |  |  |  |
|  | Non | 68.0 | 52.7 | 48.0 | 46.8 | 45.8 |
|  | Mild to moderate | 23.5 | 35.9 | 38.7 | 39.9 | 41.4 |
|  | Heavy | 8.5 | 11.3 | 13.3 | 13.3 | 12.8 |
| Physical activity | |  |  |  |  |  |
|  | Never | 63.3 | 52.4 | 51.2 | 48.1 | 40.7 |
|  | Irregular | 20.5 | 28.3 | 29.9 | 32.1 | 35.5 |
|  | Regular† | 16.1 | 19.4 | 19.0 | 19.9 | 23.7 |
| Body mass index, kg/m^2^ | |  |  |  |  |  |
|  | <18.5 | 3.2 | 1.5 | 1.5 | 1.2 | 0.8 |
|  | 18.5-<23 | 27.1 | 25.5 | 25.2 | 23.4 | 21.8 |
|  | 23-<25 | 20.4 | 24.3 | 24.1 | 24.5 | 25.8 |
|  | 25-<30 | 37.1 | 40.5 | 40.7 | 42.4 | 44.2 |
|  | ≥30 | 12.3 | 8.2 | 8.6 | 8.5 | 7.4 |
| Abdominal obesity, yes | | 40.7 | 35.0 | 34.2 | 35.4 | 35.4 |
| Hypertension, yes | | 54.6 | 47.9 | 46.2 | 45.4 | 45.5 |
| Dyslipidemia, yes | | 50.6 | 36.8 | 35.5 | 36.1 | 38.8 |
| Chronic kidney disease, yes | | 9.0 | 6.1 | 5.5 | 5.9 | 7.2 |
| Depression, yes | | 17.1 | 3.9 | 3.7 | 3.6 | 3.6 |
| Use of antidiabetic medication (n≥3) | | 23.0 | 12.1 | 11.7 | 11.5 | 11.8 |
| Insulin treatment, yes | | 16.5 | 5.7 | 5.7 | 5.5 | 5.6 |
| Type 2 diabetes duration | |  |  |  |  |  |
|  | Newly diagnosed | 25.3 | 53.7 | 55.7 | 55.7 | 51.6 |
|  | <5 years | 33.6 | 23.2 | 22.5 | 22.4 | 24.0 |
|  | ≥5 years | 41.1 | 23.2 | 21.7 | 21.9 | 24.4 |
| **Mean (SD)** | |  |  |  |  |  |
| Age, y | | 52.0 (7.8) | 52.2 (8.3) | 50.6 (8.9) | 50.4 (9.1) | 51.5 (7.9) |
| Body mass index, kg/m^2^ | | 25.2 (4.2) | 25.1 (3.5) | 25.2 (3.5) | 25.3 (3.4) | 25.3 (3.2) |
| Waist circumference, cm | | 85.4 (10.1) | 84.7 (8.8) | 84.8 (8.8) | 85.3 (8.5) | 85.6 (8.3) |
| Fasting glucose, mg/dL | | 146 (56) | 142 (52) | 142 (52) | 140 (50) | 139 (46) |
| Blood pressure, mm Hg | |  |  |  |  |  |
|  | Systolic | 125 (16) | 128 (16) | 128 (16) | 128 (15) | 127 (15) |
|  | Diastolic | 78 (10) | 80 (10) | 80 (10) | 80 (10) | 79 (10) |
| Total cholesterol, mg/dL | | 191 (44) | 201 (42) | 201 (42) | 201 (42) | 199 (41) |

Data are presented as percentages for categorical variables and means (standard deviation) for continuous variables.

*Quartile 1 (Q1) is the lowest income category, whereas Quartile 4 (Q4) is the highest. Medical Aids beneficiaries, included as a part of Quartile 1 elsewhere, were categorized separately.

†Regular exercise was defined to be at least 30 minutes of moderate physical activity for ≥5 days weekly or at least 20 minutes of strenuous physical activity ≥3 days weekly

**Supplementary table 3. Distribution of cumulative number of years being in low- or high-income group according to income variability quartile**

|  | **Income variability quartile** | | | | |
| --- | --- | --- | --- | --- | --- |
|  | Quartile 1 | Quartile 2 | Quartile 3 | Quartile 4 | *P* value |
| N | 381,427 | 382,586 | 381,060 | 383,035 |  |
| **Cumulative number of years being in low-income group** | | | |  | <.0001 |
| 0 | 350,858 (91.99) | 355,437 (92.9) | 212,445 (55.75) | 22,856 (5.97) |  |
| 1 | 117 (0.03) | 7,300 (1.91) | 55,197 (14.49) | 123,747 (32.31) |  |
| 2 | 95 (0.02) | 5,058 (1.32) | 31,597 (8.29) | 85,485 (22.32) |  |
| 3 | 27 (0.01) | 3,216 (0.84) | 20,780 (5.45) | 66,470 (17.35) |  |
| 4 | NA | 2,717 (0.71) | 16,793 (4.41) | 53,625 (14.00) |  |
| 5 | 30,330 (7.95) | 8,858 (2.32) | 44,248 (11.61) | 30,852 (8.05) |  |
| **Cumulative number of years being in high-income group** | | | |  | <.0001 |
| 0 | 103,987 (27.26) | 225,227 (58.87) | 261,357 (68.59) | 249,110 (65.04) |  |
| 1 | 8,895 (2.33) | 29,243 (7.64) | 41,362 (10.85) | 55,858 (14.58) |  |
| 2 | 9,062 (2.38) | 25,891 (6.77) | 30,027 (7.88) | 34,872 (9.10) |  |
| 3 | 14,101 (3.70) | 27,997 (7.32) | 25,376 (6.66) | 24,464 (6.39) |  |
| 4 | 13,481 (3.53) | 33,500 (8.76) | 22,882 (6.00) | 18,731 (4.89) |  |
| 5 | 231,901 (60.80) | 40,728 (10.65) | 56 (0.01) | NA |  |

Quartile 1 is the lowest category of income variability, whereas Quartile 4 is the highest.

**Supplementary table 4. Association between various indicators of income dynamics and risk of composite cardiovascular events in individuals with type 2 diabetes, with further adjustment for the potential mediators such as obesity and lifestyle factors, as well as initial income quantiles in the preceding 4 years**

|  |  | Number of subjects | Number of events | Total number of person-years of follow-up | Incidence rate  (per 1,000 person-years) | Model 4 | Model 5 |
| --- | --- | --- | --- | --- | --- | --- | --- |
|  |  |  |  |  |  | HR (95% CI) | HR (95% CI) |
| **Cumulative number of years being in low-income group** | | | | | | | |
|  | 0 | 941,596 | 61,936 | 6,902,799 | 8.97 | 1 (Ref.) | 1 (Ref.) |
|  | 1 | 186,361 | 13,894 | 1,340,883 | 10.36 | 1.14 (1.12, 1.16) | 1.09 (1.07, 1.11) |
|  | 2 | 122,235 | 9,296 | 882,994 | 10.53 | 1.16 (1.13, 1.18) | 1.10 (1.07, 1.13) |
|  | 3 | 90,493 | 7,323 | 651,329 | 11.24 | 1.21 (1.18, 1.24) | 1.14 (1.11, 1.17) |
|  | 4 | 73,135 | 6,190 | 526,123 | 11.77 | 1.23 (1.20, 1.26) | 1.15 (1.11, 1.18) |
|  | 5 | 114,288 | 10,680 | 802,853 | 13.30 | 1.33 (1.31, 1.36) | 1.19 (1.15, 1.22) |
|  | *P for trend* |  |  |  |  | <.0001 | <.0001 |
| **Cumulative number of years being in high-income group** | | | | | | | |
|  | 0 | 839,681 | 65,191 | 6,075,584 | 10.73 | 1 (Ref.) | 1 (Ref.) |
|  | 1 | 135,358 | 9,675 | 980,839 | 9.86 | 0.89 (0.88, 0.91) | 0.93 (0.91, 0.95) |
|  | 2 | 99,852 | 6,836 | 726,889 | 9.40 | 0.85 (0.83, 0.87) | 0.89 (0.86, 0.91) |
|  | 3 | 91,938 | 5,930 | 672,727 | 8.81 | 0.80 (0.78, 0.82) | 0.85 (0.82, 0.87) |
|  | 4 | 88,594 | 5,691 | 642,598 | 8.86 | 0.81 (0.78, 0.83) | 0.85 (0.83, 0.88) |
|  | 5 | 272,685 | 15,996 | 2,008,344 | 7.96 | 0.74 (0.73, 0.75) | 0.81 (0.78, 0.83) |
|  | *P for trend* |  |  |  |  | <.0001 | <.0001 |
| **Income variability quartile*** | | | | | | | |
|  | Quartile 1 | 381,427 | 24,416 | 2,793,965 | 8.74 | 1 (Ref.) | 1 (Ref.) |
|  | Quartile 2 | 382,586 | 26,163 | 2,809,034 | 9.31 | 1.06 (1.04, 1.08) | 0.99 (0.98, 1.01) |
|  | Quartile 3 | 381,060 | 28,514 | 2,757,206 | 10.34 | 1.14 (1.12, 1.16) | 1.04 (1.02, 1.06) |
|  | Quartile 4 | 383,035 | 30,226 | 2,746,777 | 11.00 | 1.20 (1.18, 1.23) | 1.08 (1.06, 1.10) |
|  | *P for trend* |  |  |  |  | <.0001 | <.0001 |
| **Baseline income status**† | | | | | | | |
|  | Medical Aids | 15,165 | 1,877 | 79,327 | 23.66 | 2.69 (2.57, 2.82) | 1.54 (1.44, 1.64) |
|  | Quartile 1 | 308,520 | 25,473 | 2,223,969 | 11.45 | 1.29 (1.27, 1.31) | 1.15 (1.13, 1.17) |
|  | Quartile 2 | 318,838 | 23,728 | 2,304,582 | 10.30 | 1.23 (1.21, 1.25) | 1.10 (1.08, 1.12) |
|  | Quartile 3 | 414,685 | 29,289 | 3,037,440 | 9.64 | 1.17 (1.15, 1.19) | 1.07 (1.06, 1.09) |
|  | Quartile 4 | 470,900 | 28,952 | 3,461,665 | 8.36 | 1 (Ref.) | 1 (Ref.) |
|  | *P for trend* |  |  |  |  | <.0001 | <.0001 |

Abbreviations: HR, hazard ratio; CI, confidence interval.

*Quartile 1 is the lowest category of income variability, whereas Quartile 4 is the highest

†Quartile 1 is the lowest income category, whereas Quartile 4 is the highest. Medical Aids beneficiaries, included as a part of Quartile 1 elsewhere, were categorized separately.

**Model 4**: adjusted for age at baseline (continuous), sex, and residential location (metropolitan, urban, or rural), depression, chronic kidney disease, hypertension, dyslipidemia, systolic blood pressure (continuous), total cholesterol (continuous), HDL-cholesterol (continuous), blood glucose concentration (continuous), number of prescriptions for oral anti-diabetic medications per year (<3, ≥3), history of insulin prescription, pack-years of smoking (never, <10, 10 to 19, or ≥20 years), alcohol consumption (never, mild to moderate, or heavy), physical activity (non, irregular, or regular), BMI (kg/m^2^, <18.5, 18.5-<23, 23-<25, 25-<30, ≥30), and the presence or absence of abdominal obesity. **Model 5**: adjusted for covariates in Model 4 plus 20 initial income quantiles at the preceding 4 years

**Supplementary table 5.** **Association between various indicators of income dynamics and the corresponding risk of individual cardiovascular events in adults with type 2 diabetes**

|  |  | **Number of subjects** | **Myocardial infarction** | | |  | **Ischemic stroke** | | |  | **Heart failure** | | |  | **Cardiovascular mortality** | | |
| --- | --- | --- | --- | --- | --- | --- | --- | --- | --- | --- | --- | --- | --- | --- | --- | --- | --- |
|  |  |  | No. of events | Incidence rate (per 1,000 person-years) | Adjusted HR (95% CI)* |  | No. of events | Incidence rate (per 1,000 person-years) | Adjusted HR (95% CI)* |  | No. of events | Incidence rate (per 1,000 person-years) | Adjusted HR (95% CI)* |  | No. of events | Incidence rate (per 1,000 person-years) | Adjusted HR (95% CI)* |
| **Cumulative number of years being in low-income group**† | | | | | | | | | | | | | | | | | |
| 0 | | 941,596 | 17,048 | 2.43 | 1 (Ref.) |  | 20,436 | 2.92 | 1 (Ref.) |  | 31,170 | 4.45 | 1 (Ref.) |  | 4,307 | 0.61 | 1 (Ref.) |
| 1 | | 186,361 | 3,727 | 2.73 | 1.15  (1.11, 1.19) |  | 4,665 | 3.43 | 1.16  (1.13, 1.20) |  | 7,160 | 5.25 | 1.19  (1.16, 1.22) |  | 1,076 | 0.78 | 1.30  (1.21, 1.39) |
| 2 | | 122,235 | 2,459 | 2.73 | 1.16  (1.11, 1.21) |  | 3,086 | 3.44 | 1.17  (1.12, 1.21) |  | 4,848 | 5.40 | 1.22  (1.18, 1.26) |  | 735 | 0.81 | 1.35  (1.25, 1.46) |
| 3 | | 90,493 | 1,910 | 2.87 | 1.20  (1.15, 1.26) |  | 2,547 | 3.85 | 1.28  (1.22, 1.33) |  | 3,769 | 5.69 | 1.26  (1.22, 1.30) |  | 622 | 0.93 | 1.51  (1.39, 1.64) |
| 4 | | 73,135 | 1,630 | 3.04 | 1.24  (1.18, 1.31) |  | 2,111 | 3.95 | 1.27  (1.21, 1.33) |  | 3,162 | 5.91 | 1.27  (1.23, 1.32) |  | 598 | 1.10 | 1.74  (1.59, 1.89) |
| 5 | | 114,288 | 2,714 | 3.31 | 1.31  (1.26, 1.36) |  | 3,670 | 4.49 | 1.38  (1.34, 1.43) |  | 5,481 | 6.70 | 1.41  (1.37, 1.45) |  | 1,004 | 1.21 | 1.81  (1.69, 1.94) |
|  | *P* for trend | |  |  | <.0001 |  |  |  | <.0001 |  |  |  | <.0001 |  |  |  | <.0001 |
| **Cumulative number of years being in high-income group**† | | | | | | | | | | | | | | | | | |
| 0 | | 839,681 | 17,260 | 2.79 | 1 (Ref.) |  | 21,938 | 3.55 | 1 (Ref.) |  | 33,253 | 5.38 | 1 (Ref.) |  | 5,497 | 0.88 | 1 (Ref.) |
| 1 | | 135,358 | 2,639 | 2.64 | 0.92  (0.88, 0.95) |  | 3,210 | 3.23 | 0.87  (0.84, 0.90) |  | 4,910 | 4.93 | 0.87  (0.85, 0.90) |  | 718 | 0.71 | 0.78  (0.72, 0.84) |
| 2 | | 99,852 | 1,824 | 2.47 | 0.84 (0.80,0.89) |  | 2,323 | 3.15 | 0.84  (0.81, 0.88) |  | 3,493 | 4.73 | 0.83  (0.80, 0.86) |  | 454 | 0.61 | 0.66  (0.60, 0.73) |
| 3 | | 91,938 | 1,633 | 2.39 | 0.82  (0.78, 0.86) |  | 1,987 | 2.91 | 0.79  (0.75, 0.83) |  | 3,027 | 4.44 | 0.78  (0.75, 0.81) |  | 383 | 0.56 | 0.61  (0.55, 0.67) |
| 4 | | 88,594 | 1,584 | 2.43 | 0.83  (0.79, 0.87) |  | 1,844 | 2.83 | 0.76  (0.73, 0.80) |  | 2,928 | 4.49 | 0.79  (0.76, 0.82) |  | 356 | 0.54 | 0.59  (0.53, 0.66) |
| 5 | | 272,685 | 4,548 | 2.23 | 0.75  (0.73, 0.78) |  | 5,213 | 2.56 | 0.71  (0.69, 0.73) |  | 7,979 | 3.92 | 0.69  (0.68, 0.71) |  | 934 | 0.46 | 0.49  (0.46, 0.53) |
|  | *P* for trend | |  |  | <.0001 |  |  |  | <.0001 |  |  |  | <.0001 |  |  |  | <.0001 |
| **Income variability quartile**‡ | | | | | | | | | | | | | | | | | |
| Q1 | | 381,427 | 6,693 | 2.36 | 1 (Ref.) |  | 8,012 | 2.83 | 1 (Ref.) |  | 12,241 | 4.32 | 1 (Ref.) |  | 1,695 | 0.59 | 1 (Ref.) |
| Q2 | | 382,586 | 7,131 | 2.50 | 1.09  (1.05, 1.12) |  | 8,649 | 3.04 | 1.08  (1.05, 1.11) |  | 13,267 | 4.65 | 1.09 (1.06,1.12) |  | 1,867 | 0.65 | 1.11  (1.04, 1.19) |
| Q3 | | 381,060 | 7,664 | 2.73 | 1.18  (1.14, 1.22) |  | 9,525 | 3.40 | 1.17  (1.13, 1.20) |  | 14,606 | 5.21 | 1.20 (1.17,1.22) |  | 2,299 | 0.81 | 1.35  (1.27, 1.44) |
| Q4 | | 383,035 | 8,000 | 2.86 | 1.24  (1.20, 1.28) |  | 10,329 | 3.70 | 1.26  (1.22, 1.30) |  | 15,476 | 5.54 | 1.26 (1.23,1.29) |  | 2,481 | 0.88 | 1.47  (1.38, 1.57) |
|  | *P* for trend | |  |  | <.0001 |  |  |  | <.0001 |  |  |  | <.0001 |  |  |  | <.0001 |
| **Baseline income status§** | | | | | | | | | | | | | | | | | |
| MA | | 15,165 | 433 | 5.29 | 2.32  (2.10, 2.55) |  | 562 | 6.90 | 2.40  (2.21, 2.62) |  | 1,063 | 13.12 | 3.81  (3.58, 4.06) |  | 190 | 2.30 | 4.70  (4.04, 5.47) |
| Q1 | | 308,520 | 6,711 | 2.96 | 1.29  (1.25, 1.33) |  | 8,795 | 3.89 | 1.37  (1.33, 1.41) |  | 12,943 | 5.72 | 1.33  (1.30, 1.36) |  | 2,242 | 0.98 | 1.88  (1.77, 2.01) |
| Q2 | | 318,838 | 6,305 | 2.69 | 1.22  (1.18, 1.26) |  | 7,999 | 3.42 | 1.29  (1.25, 1.33) |  | 12,052 | 5.15 | 1.27  (1.24, 1.31) |  | 1,963 | 0.83 | 1.70  (1.60, 1.82) |
| Q3 | | 414,685 | 7,944 | 2.57 | 1.16  (1.13, 1.20) |  | 9,762 | 3.17 | 1.21  (1.18, 1.25) |  | 14,794 | 4.80 | 1.19  (1.16, 1.21) |  | 2,191 | 0.70 | 1.47  (1.38, 1.56) |
| Q4 | | 470,900 | 8,095 | 2.30 | 1 (Ref.) |  | 9,397 | 2.68 | 1 (Ref.) |  | 14,738 | 4.20 | 1 (Ref.) |  | 1,756 | 0.50 | 1 (Ref.) |
|  | *P* for trend | |  |  | <.0001 |  |  |  | <.0001 |  |  |  | <.0001 |  |  |  | <.0001 |

Abbreviations: HR, hazard ratio; CI, confidence interval; Q, quartile; MA, medical aids.

*Adjusted for age at baseline (continuous), sex, residential location (metropolitan, urban, or rural), depression, chronic kidney disease, hypertension, dyslipidemia, systolic blood pressure (continuous), total cholesterol (continuous), HDL-cholesterol (continuous), blood glucose concentration (continuous), number of prescriptions for oral anti-diabetic medications per year (<3, ≥3), and history of insulin prescription.

†The number of times an individual was categorized in the low- or high-income group was counted every year during the 4 years prior to the baseline year of 2009 through 2012

‡Quartile 1 (Q1) is the lowest category of income variability, whereas Quartile 4 (Q4) is the highest.

§Quartile 1 (Q1) is the lowest income category, whereas Quartile 4 (Q4) is the highest. Medical Aids beneficiaries, included as a part of Quartile 1 elsewhere, were categorized separately.

**Supplementary table 6. Changes in income status between the two time points (4 years ago vs. baseline) and the corresponding risk of composite cardiovascular events in individuals with type 2 diabetes, with further adjustment for the potential mediators such as obesity and lifestyle factors, as well as initial income quantiles in the preceding 4 years**

| **Income status** | | Number of subjects | Number of events | Total number of person-years of follow-up | Incidence rate (per 1,000 person-years) | Model 4 | Model 5 |
| --- | --- | --- | --- | --- | --- | --- | --- |
| **First time point -**  preceding 4 years (2004-2007) | **Last time point** -  baseline  (2009-2012) |  |  |  |  |  |  |
|  |  |  |  |  |  | HR (95% CI) | HR (95% CI) |
| Medical Aids | Medical Aids | 11,218 | 1,374 | 57,840 | 23.76 | 1 (Ref.) | 1 (Ref.) |
|  | Quartile 1 | 5,738 | 688 | 37,341 | 18.42 | 0.72 (0.66, 0.79) | 0.86 (0.79, 0.95) |
|  | Quartile 2 | 4,084 | 460 | 27,217 | 16.90 | 0.70 (0.63, 0.78) | 0.85 (0.77, 0.95) |
|  | Quartile 3 | 2,067 | 265 | 13,877 | 19.10 | 0.75 (0.66, 0.85) | 0.92 (0.80, 1.05) |
|  | Quartile 4 | 470 | 61 | 3,184 | 19.16 | 0.73 (0.56, 0.94) | 0.90 (0.70, 1.17) |
|  | P for trend |  |  |  |  | <.0001 | <.0001 |
| Quartile 1 | Medical Aids | 1,784 | 259 | 9,496 | 27.27 | 2.45 (2.17, 2.77) | 1.97 (1.74, 2.23) |
|  | Quartile 1 | 141,786 | 12,206 | 1,030,569 | 11.84 | 1 (Ref.) | 1 (Ref.) |
|  | Quartile 2 | 82,197 | 6,123 | 600,869 | 10.19 | 0.97 (0.94, 1.00) | 0.97 (0.94, 1.00) |
|  | Quartile 3 | 46,686 | 3,547 | 339,805 | 10.44 | 0.96 (0.93, 1.00) | 0.95 (0.92, 0.99) |
|  | Quartile 4 | 20,004 | 1,591 | 143,850 | 11.06 | 0.90 (0.86, 0.95) | 0.89 (0.85, 0.94) |
|  | *P for trend* |  |  |  |  | <.0001 | <.0001 |
| Quartile 2 | Medical Aids | 1,392 | 168 | 7,639 | 21.99 | 2.14 (1.84, 2.50) | 1.78 (1.53, 2.08) |
|  | Quartile 1 | 63,330 | 5,355 | 450,431 | 11.89 | 1.09 (1.05, 1.12) | 1.08 (1.05, 1.12) |
|  | Quartile 2 | 126,620 | 9,501 | 912,348 | 10.41 | 1 (Ref.) | 1 (Ref.) |
|  | Quartile 3 | 108,820 | 7,674 | 799,615 | 9.60 | 0.97 (0.94, 1.00) | 0.98 (0.95, 1.01) |
|  | Quartile 4 | 25,908 | 1,873 | 186,099 | 10.06 | 0.89 (0.85, 0.94) | 0.89 (0.85, 0.94) |
|  | *P for trend* |  |  |  |  | <.0001 | <.0001 |
| Quartile 3 | Medical Aids | 579 | 55 | 3,262 | 16.86 | 1.96 (1.50, 2.55) | 1.63 (1.25, 2.13) |
|  | Quartile 1 | 56,534 | 4,370 | 407,756 | 10.72 | 1.10 (1.06, 1.13) | 1.09 (1.06, 1.13) |
|  | Quartile 2 | 67,968 | 5,037 | 489,541 | 10.29 | 1.05 (1.02, 1.08) | 1.04 (1.01, 1.08) |
|  | Quartile 3 | 196,953 | 13,520 | 1,449,896 | 9.32 | 1 (Ref.) | 1 (Ref.) |
|  | Quartile 4 | 111,424 | 6,737 | 826,919 | 8.15 | 0.91 (0.88, 0.94) | 0.92 (0.89, 0.95) |
|  | *P for trend* |  |  |  |  | <.0001 | <.0001 |
| Quartile 4 | Medical Aids | 192 | 21 | 1,090 | 19.27 | 2.63 (1.71, 4.03) | 2.15 (1.40, 3.30) |
|  | Quartile 1 | 41,132 | 2,854 | 297,873 | 9.58 | 1.12 (1.07, 1.16) | 1.10 (1.05, 1.14) |
|  | Quartile 2 | 37,969 | 2,607 | 274,607 | 9.49 | 1.10 (1.06, 1.15) | 1.08 (1.04, 1.13) |
|  | Quartile 3 | 60,159 | 4,283 | 434,247 | 9.86 | 1.10 (1.07, 1.14) | 1.07 (1.04, 1.11) |
|  | Quartile 4 | 313,094 | 18,690 | 2,301,613 | 8.12 | 1 (Ref.) | 1 (Ref.) |
|  | *P for trend* |  |  |  |  | <.0001 | <.0001 |

Abbreviations: HR, hazard ratio; CI, confidence interval.

Quartile 1 is the lowest income category, whereas Quartile 4 is the highest. Medical Aids beneficiaries, included as a part of Quartile 1 elsewhere, were categorized separately.

**Model 4**: adjusted for age at baseline (continuous), sex, and residential location (metropolitan, urban, or rural), depression, chronic kidney disease, hypertension, dyslipidemia, systolic blood pressure (continuous), total cholesterol (continuous), HDL-cholesterol (continuous), blood glucose concentration (continuous), number of prescriptions for oral anti-diabetic medications per year (<3, ≥3), history of insulin prescription, pack-years of smoking (never, <10, 10 to 19, or ≥20 years), alcohol consumption (never, mild to moderate, or heavy), physical activity (non, irregular, or regular), BMI (kg/m^2^, <18.5, 18.5-<23, 23-<25, 25-<30, ≥30), and the presence or absence of abdominal obesity. **Model 5**: adjusted for covariates in Model 4 plus 20 initial income quantiles at the preceding 4 years

**Supplementary table 7. Association between various indicators of income dynamics and risk of composite cardiovascular events in individuals with type 2 diabetes, after excluding those who had a prior cancer history**

|  | Variables | Number of subjects | Number of events | Total number of person-years of follow-up | Incidence rate  (per 1,000 person-years) | HR (95% CI)* |
| --- | --- | --- | --- | --- | --- | --- |
| Cumulative number of years being in low-income group | 0 | 923,061 | 60,438 | 6,776,047 | 8.92 | 1 (Ref.) |
|  | 1 | 182,738 | 13,542 | 1,316,577 | 10.29 | 1.16 (1.14, 1.18) |
|  | 2 | 119,846 | 9,070 | 866,867 | 10.46 | 1.18 (1.16, 1.21) |
|  | 3 | 88,693 | 7,128 | 639,293 | 11.15 | 1.24 (1.21, 1.27) |
|  | 4 | 71,705 | 6,050 | 516,507 | 11.71 | 1.26 (1.23, 1.30) |
|  | 5 | 112,113 | 10,437 | 789,177 | 13.23 | 1.38 (1.35, 1.41) |
|  | *p for trend* |  |  |  |  | <.0001 |
| Cumulative number of years being in high-income group | 0 | 825,042 | 63,755 | 5,977,916 | 10.67 | 1 (Ref.) |
|  | 1 | 132,608 | 9,448 | 962,278 | 9.82 | 0.88 (0.87, 0.90) |
|  | 2 | 97,657 | 6,615 | 712,022 | 9.29 | 0.83 (0.81, 0.85) |
|  | 3 | 89,951 | 5,772 | 659,122 | 8.76 | 0.78 (0.76, 0.80) |
|  | 4 | 86,473 | 5,517 | 628,164 | 8.78 | 0.78 (0.76, 0.81) |
|  | 5 | 266,425 | 15,558 | 1,964,966 | 7.92 | 0.71 (0.70, 0.72) |
|  | *p for trend* |  |  |  |  | <.0001 |
| Income variability quartile† | Quartile 1 | 373450 | 23,788 | 2739722 | 8.68 | 1 (Ref.) |
|  | Quartile 2 | 375768 | 25,582 | 2762456 | 9.26 | 1.09 (1.07, 1.11) |
|  | Quartile 3 | 373929 | 27,856 | 2709015 | 10.28 | 1.18 (1.16, 1.20) |
|  | Quartile 4 | 375009 | 29,439 | 2693275 | 10.93 | 1.25 (1.22, 1.27) |
|  | *p for trend* |  |  |  |  | <.0001 |
| Baseline income status‡ | Medical Aids | 14,591 | 1,799 | 76,586 | 23.49 | 2.94 (2.80, 3.09) |
|  | Quartile 1 | 302,872 | 24,924 | 2,185,814 | 11.40 | 1.34 (1.31, 1.36) |
|  | Quartile 2 | 313,246 | 23,166 | 2,267,085 | 10.22 | 1.27 (1.25, 1.29) |
|  | Quartile 3 | 407,079 | 28,631 | 2,985,793 | 9.59 | 1.20 (1.18, 1.22) |
|  | Quartile 4 | 460,368 | 28,145 | 3,389,190 | 8.30 | 1 (Ref.) |
|  | *p for trend* |  |  |  |  | <.0001 |

Abbreviations: HR, hazard ratio; CI, confidence interval.

*adjusted for age at baseline (continuous), sex, and residential location (metropolitan, urban, or rural), depression, chronic kidney disease, hypertension, dyslipidemia, systolic blood pressure (continuous), total cholesterol (continuous), HDL-cholesterol (continuous), blood glucose concentration (continuous), number of prescriptions for oral anti-diabetic medications per year (<3, ≥3), history of insulin prescription

†Quartile 1 is the lowest category of income variability whereas Quartile 4 is the highest

‡Medical Aids beneficiaries, included as a part of Quartile 1 elsewhere, were categorized separately

**Supplementary table 8. 5-year landmark analyses of the risk of composite cardiovascular events according to the cumulative number of years being in low- or high-income group, income variability quartile, and baseline income status**

|  |  | Number of subjects | Number of events | Total number of person-years of follow-up | Incidence rate  (per 1,000 person-years) | HR (95% CI)* |
| --- | --- | --- | --- | --- | --- | --- |
| Cumulative number of years being in low-income group | 0 | 906,879 | 38,930 | 3,201,765 | 12.16 | 1 (Ref.) |
|  | 1 | 178,093 | 8,469 | 611,047 | 13.86 | 1.14 (1.12, 1.17) |
|  | 2 | 116,798 | 5,835 | 404,380 | 14.43 | 1.19 (1.15, 1.22) |
|  | 3 | 86,169 | 4,564 | 297,526 | 15.34 | 1.24 (1.20, 1.28) |
|  | 4 | 69,462 | 3,802 | 240,571 | 15.80 | 1.24 (1.20, 1.29) |
|  | 5 | 107,251 | 6,213 | 358,922 | 17.31 | 1.32 (1.28, 1.36) |
|  | *p for trend* |  |  |  |  | <.0001 |
| Cumulative number of years being in high-income group | 0 | 800,482 | 40,034 | 2,790,975 | 14.34 | 1 (Ref.) |
|  | 1 | 129,896 | 6,069 | 449,663 | 13.50 | 0.90 (0.88, 0.92) |
|  | 2 | 96,052 | 4,279 | 334,595 | 12.79 | 0.84 (0.82, 0.87) |
|  | 3 | 88,579 | 3,705 | 311,282 | 11.90 | 0.79 (0.76, 0.82) |
|  | 4 | 85,462 | 3,566 | 294,131 | 12.12 | 0.80 (0.77, 0.83) |
|  | 5 | 264,181 | 10,160 | 933,566 | 10.88 | 0.72 (0.71, 0.74) |
|  | *p for trend* |  |  |  |  | <.0001 |
| Income variability quartile† | Quartile 1 | 367,643 | 15,197 | 2,739,722 | 11.74 | 1 (Ref.) |
|  | Quartile 2 | 367,671 | 16,361 | 2,762,456 | 12.52 | 1.09 (1.06, 1.11) |
|  | Quartile 3 | 364,266 | 17,650 | 2,709,015 | 13.96 | 1.18 (1.15, 1.20) |
|  | Quartile 4 | 365,072 | 18,605 | 2,693,275 | 14.90 | 1.24 (1.22, 1.27) |
|  | *p for trend* |  |  |  |  | <.0001 |
| Baseline income status‡ | Medical Aids | 13,388 | 759 | 22,002 | 34.50 | 3.24 (3.01, 3.49) |
|  | Quartile 1 | 293,420 | 15,709 | 1,018,480 | 15.42 | 1.31 (1.28, 1.34) |
|  | Quartile 2 | 304,584 | 14,693 | 1,056,133 | 13.91 | 1.25 (1.23, 1.28) |
|  | Quartile 3 | 397,977 | 18,291 | 1,410,175 | 12.97 | 1.18 (1.16, 1.21) |
|  | Quartile 4 | 455,283 | 18,361 | 1,607,422 | 11.42 | 1 (Ref.) |
|  | *p for trend* |  |  |  |  | <.0001 |

Abbreviations: HR, hazard ratio; CI, confidence interval.

*adjusted for age at baseline (continuous), sex, and residential location (metropolitan, urban, or rural), depression, chronic kidney disease, hypertension, dyslipidemia, systolic blood pressure (continuous), total cholesterol (continuous), HDL-cholesterol (continuous), blood glucose concentration (continuous), number of prescriptions for oral anti-diabetic medications per year (<3, ≥3), history of insulin prescription.

†Quartile 1 is the lowest category of income variability, whereas Quartile 4 is the highest.

‡Quartile 1 is the lowest income category, whereas Quartile 4 is the highest. Medical Aids beneficiaries, included as a part of Quartile 1 elsewhere, were categorized separately.

**Supplementary figure 1. Change of income status between the two-time points (4 years ago vs. baseline) and the corresponding risk of individual cardiovascular events in individuals with type 2 diabetes: (A) myocardial infarction, (B) ischemic stroke, (C) heart failure, and (D) cardiovascular mortality**

(A) Myocardial infarction

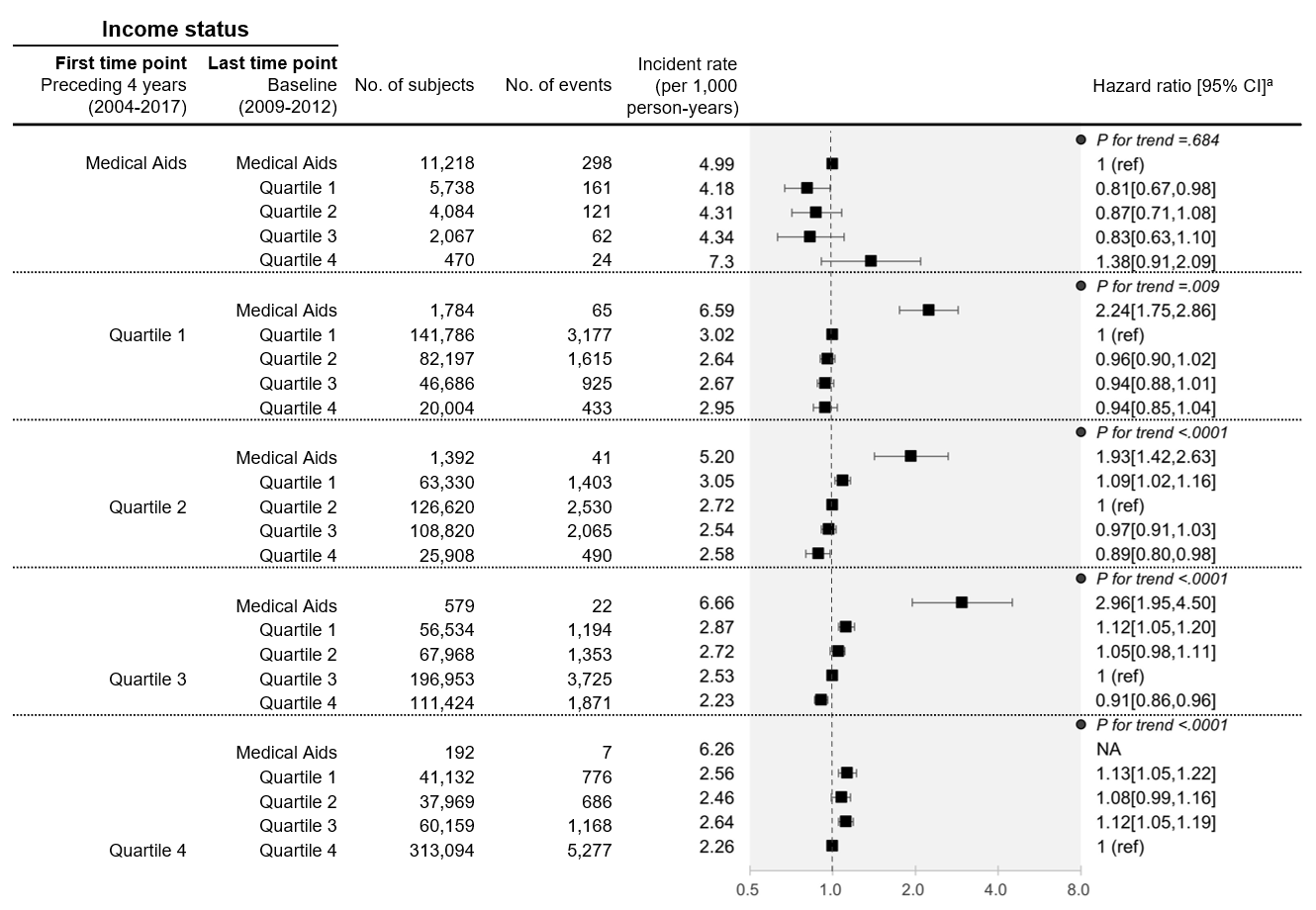

(B) Ischemic stroke

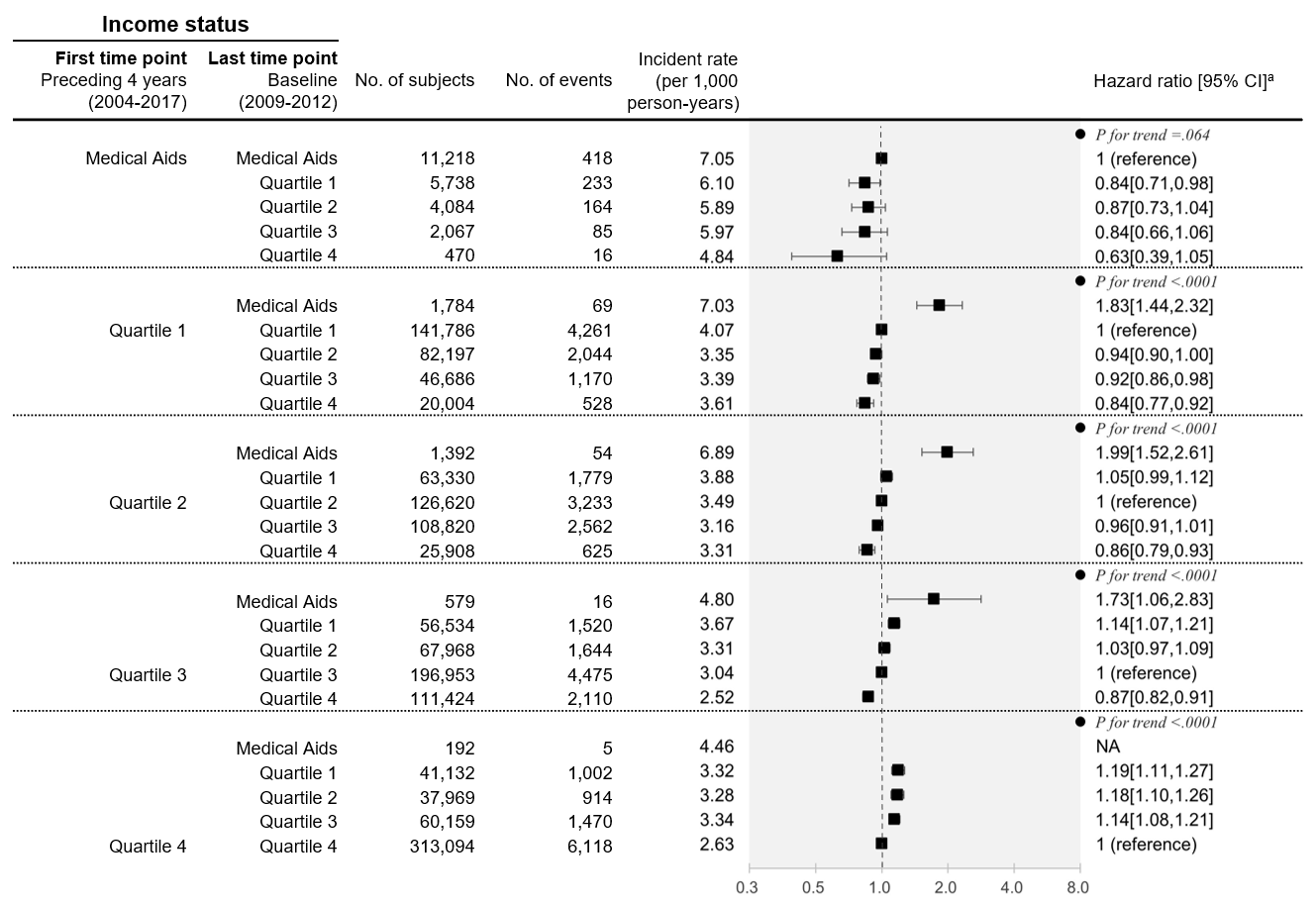

(C) Heart failure

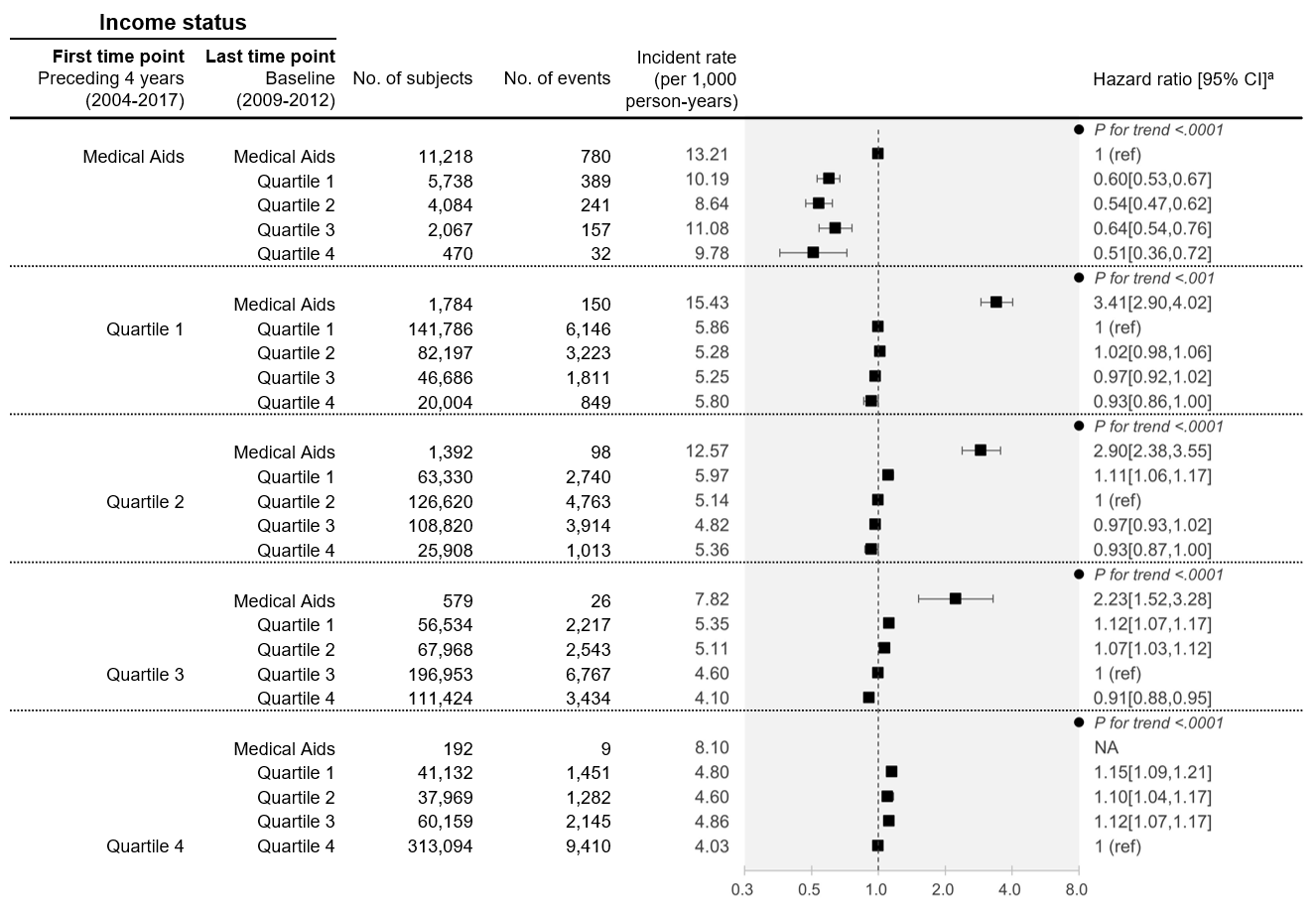

(D) Cardiovascular mortality

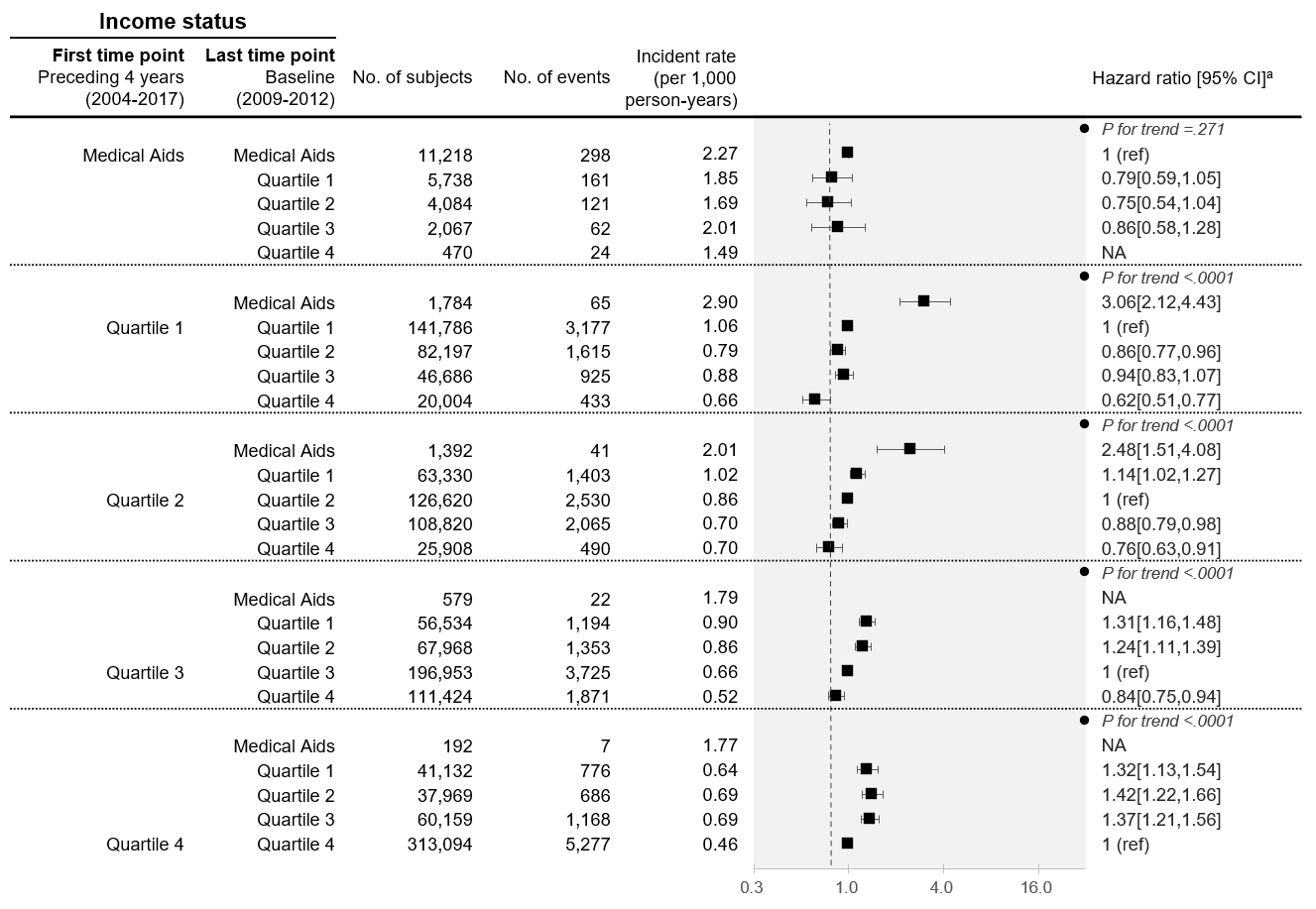

Quartile 1 is the lowest income category, whereas Quartile 4 is the highest. Medical Aids beneficiaries, included as a part of Quartile 1 elsewhere, were categorized separately.

NA, number of events were less than 10 cases.

*adjusted for age at baseline (continuous), sex, and residential location (metropolitan, urban, or rural), depression, chronic kidney disease, hypertension, dyslipidemia, systolic blood pressure (continuous), total cholesterol (continuous), HDL-cholesterol (continuous), blood glucose concentration (continuous), number of prescriptions for oral anti-diabetic medications per year (<3, ≥3), history of insulin prescription.

**Supplementary figure 2.** **Association between various indicators of income dynamics and risk of composite cardiovascular events in individuals with type 2 diabetes stratified by selected factors**

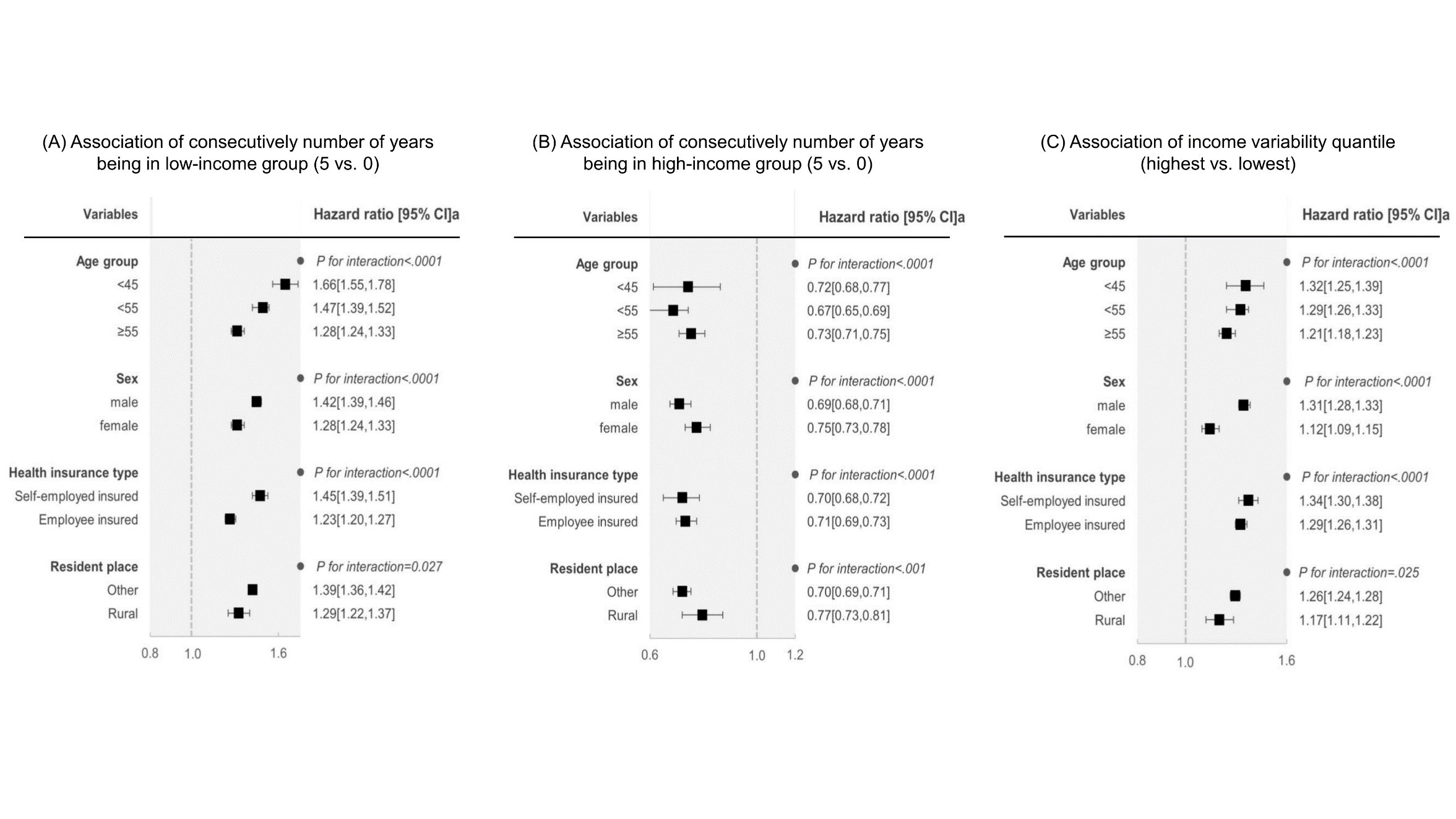

*adjusted for age at baseline (continuous), sex, and residential location (metropolitan, urban, or rural), depression, chronic kidney disease, hypertension, dyslipidemia, systolic blood pressure (continuous), total cholesterol (continuous), HDL-cholesterol (continuous), blood glucose concentration (continuous), number of prescriptions for oral anti-diabetic medications per year (<3, ≥3), history of insulin prescription except a stratifying variable
